# Young adults vulnerability to COVID-19 in Brazil: an overview across the country

**DOI:** 10.1101/2021.09.27.21264086

**Authors:** Fernanda Sumika Hojo de Souza, Natália Satchiko Hojo-Souza, Daniela Carine Ramires de Oliveira, Cristiano Maciel da Silva, Daniel Ludovico Guidoni

## Abstract

Brazil is a country of continental dimensions, where many smaller countries would fit. In addition to demographic, socioeconomic, and cultural differences, hospital infrastructure and healthcare varies across all 27 federative units. Therefore, the evolution of COVID-19 pandemic did not manifest itself in a homogeneous and predictable trend across the nation. In late 2020 and early 2021, new waves of the COVID-19 outbreak have caused an unprecedented sanitary collapse in Brazil. Unlike the first COVID-19 wave, in subsequent waves, preliminary evidence has pointed to an increase in the daily reported cases among younger people being hospitalized, overloading the healthcare system. In this comprehensive retrospective study, confirmed cases of hospitalization, ICU admission, IMV requirement and in-hospital death from Brazilian COVID-19 patients throughout 2020 until the beginning of 2021 were analyzed through a spatio-temporal study for patients aged 20-59 years. All Brazilian federative units had their data disaggregated in six periods of ten epidemiological weeks each. We found that there is a wide variation in the waves dynamic due to SARS-CoV-2 infection, both in the first and in subsequent outbreaks in different federative units over the analyzed periods. As a result, atypical waves can be seen in the Brazil data as a whole. The analysis showed that Brazil is experiencing a numerical explosion of hospitalizations and deaths for patients aged 20-59 years, especially in the state of São Paulo, with a similar proportion of hospitalizations for this age group but higher proportion of deaths compared to the first wave.

## Introduction

COVID-19, a highly infectious disease caused by the SARS-CoV-2 virus, has become an unprecedented global threat in the contemporary history of the worldwide population. The initial epidemic in Wuhan (China), in December 2019, spread and reached most nations, becoming a pandemic of difficult containment^1^. Many countries seriously affected by the COVID-19 pandemic are facing a second wave due to the continued transmission of SARS-CoV-2. Like many countries, Brazil experienced a first COVID-19 wave that started in March 2020. The first COVID-19 cases in Brazil emerged in Amazonas and São Paulo, two states that host important international airports. After the first SARS-CoV-2 dissemination wave across the country, continuous transmission generated subsequent waves that, so far, have not been contained.

In Brazil, testing of infected people and contact tracking have been poorly implemented, making “Non-Pharmaceutical Interventions” (NPIs) the main measures for to mitigate or suppress transmission during the first wave. However, prolonged NPIs have a direct impact on economic and social dimensions of the population, leading to reduced adherence to measures and causing the COVID-19 recrudescence.

Another factor that may contribute to the occurrence of new outbreaks is the SARS-CoV-2 variants emergence. Indeed, the emergence of the second wave in the United Kingdom (UK) was due to “Variant of Concern” (VOC) 202012/01 (lineage B.1.1.7), detected in September 2020, which is more transmissible^2^ and causes more severe illness^3^. This variant became the dominant lineage in the UK and has spread to many countries. In Brazil, a new SARS-CoV-2 variant B.1.1.28.1 (named P.1) was detected in the Amazonas State on January 12, 2021, being classified as VOC due to several important mutations and higher transmissibility^4^,^5^.

P.1 quickly became the dominant variant and probably responsible for the second wave that emerged in mid-December 2020 in Amazonas^5^. Recent study on the detection of VOCs through a mutation common to three variants (P.1 in the Amazonas, B.1.1.7, in the United Kingdom and B.1.351, in South Africa) found the presence of the variant in several Brazilian states, possibly with a prevalence of P.1^6^.

Brazil is a country of continental dimensions, with a total area of 8,514,877 km^27^, divided into 27 federative units (26 states and the Federal District) (Figure S2). With these characteristics, Brazil has showed significant differences regarding to demographic, socioeconomic and cultural factors. In addition, hospital infrastructure and healthcare is varied across all 27 federative units, despite the existence of the Unified Health System (Sistema Único de Saúde - SUS)^8^ which guarantees full, universal and free access for the Brazilian population. In this context, the evolution of COVID-19 pandemic did not manifest itself in a homogeneous and predictable trend across the nation^9^. Studies covering the first COVID-19 wave showed that the disease affects the Brazilian population unevenly, with the mortality of hospitalized COVID-19 patients being higher in the North and Northeast region^10,11^.

Extensive literature has shown that older age and to have comorbidities, such as heart disease, diabetes, chronic lung disease and obesity contribute to a more severe disease outcome^12–14^. We also observed the same in a comprehensive retrospective study involving patients affected by COVID-19 during the first wave in Brazil^11^. Importantly, elderly represent the population at high lethality risk, while, younger people account for the majority population with the greatest potential for transmission of SARS-CoV-2. According to IBGE^15^, the working-age population consisting of young adults (15-64 years old) is 69.3% in Brazil (Figure S3). Unlike the first COVID-19 wave in Brazil, in subsequent waves, preliminary evidence has pointed to an increase in the cases number of younger people being hospitalized, overloading the healthcare system^16^. Factors such as reduced adherence to non-pharmaceutical interventions and the emergence of the new variant P.1, more transmissible, are possibly contributing to the current uncontrolled outbreaks in Brazil and changing the age profile of hospitalized patients.

In a preliminary analysis, we observed that in the second COVID-19 wave in the Amazonas state there was an increase in the total number of hospitalizations, Intensive Care Unit (ICU) admission, Invasive Mechanical Ventilation (IMV) requirement and in-hospital deaths compared to the first wave. In addition, there was a significant increase in ICU admission, IMV need and mortality of patients aged 20-59 years old^17^. The second outbreak coincided with the P.1 variant emergence, which was not successfully contained and is now being detected in samples from patients in several Brazilian states. Thus, a spatio-temporal analysis can reveal the nationwide scenario and contribute to the implementation of more restrictive and efficient measures where there is greater vulnerability while there is a COVID-19 vaccines shortage.

## Methods

### Data

We developed a retrospective cohort study analyzing data from epidemiological week 2020-8 to 2021-14, reported in SIVEP-Gripe^18^ (Influenza Epidemiological Surveillance Information System maintained by the Ministry of Health of Brazil). Data was accessed on May 13th, 2021. The Brazilian Severe Acute Respiratory Illness (SARI) database contains individual information of patients hospitalized due to SARI caused by different agents as SARS-CoV-2, Influenza, Adenovirus, among others. The data was extracted to select COVID-19 patients confirmed through laboratorial tests performed by molecular (RT-PCR) or immunological diagnostic (screening for antibodies or antigens).

Information regarding young adults (age group 20-59 years old) for hospitalization (*n* = 507, 631), ICU admission (*n* = 147, 195), IMV requirement (*n* = 74, 580) and in-hospital death (*n* = 96, 994) in the 27 federative units of Brazil was retrieved. It is important to highlight that until week 2021-14, there was no vaccination for 20-59 years, even those with comorbidities. By April 10th, 2021 (week 2021-14), only 11% of the population had received the first dose and 3.3% the second dose of vaccine in Brazil, giving priority to the elderly and health professionals^19^. Figure S1 presents a diagram of SIVEP-Gripe data used in this study.

### Variables and statistical analysis

The variables used in this study are classified accordingly: categorical binary variables (cure/death, ICU admission, IMV requirement), categorical ordinal variable (age group), and categorical nominal variable (Brazilian federative unit). Descriptive statistics are used to provide the features of the data in study.

Aiming to reveal the dynamics of COVID-19, we have analyzed data by six periods of 10 weeks, named T1 to T6. Periods comprised the epidemiological weeks (EW) of the onset of symptoms in the patients as follows. T1: EW 2020-8 to 2020-17 (February, 16th to April, 25th), T2: EW 2020-18 to 2020-27 (April, 26th to July, 4th), T3: EW 2020-28 to 2020-37 (July, 5th to September, 12th), T4: EW 2020-38 to 2020-47 (September, 13th to November, 21st), T5: EW 2020-48 to 2021-4 (November, 22nd to January, 30th) and T6: EW 2021-5 to 2021-14 (January, 31st to April 10th).

Besides absolute numbers, we used proportions to analyze the data. The hospitalization percentage of patients aged 20-59 is given considering all other age groups. The other proportions are given specifically for the 20-59 age group. The proportion of patients requiring ICU and IMV were calculated based on the total patients owning such information in the database. Therefore, each variable presents a different *n* value.

In-hospital Case Fatality Rate (hCFR) is given as the number of deaths by the number of hospitalized patients with closed outcome (cure or death) in the given period.

In order to compare the studied periods, we have calculated a test of statistical significance (chi-square)^20^ and Risk Ratios (RR) with 95% Confidence Interval (CI) to measure the strength of association^21^. The Pearson correlation coefficient was used to compare two variables along the time series data. All analyses were performed using `Python` (version 3.7.10), the statistical package `scipy` (version 1.4.1) and the epidemiological package `epipy` (version 0.0.2.1). *P*-values < 0.05 were considered statistically significant.

### Ethics statement

This retrospective study is based on a publicly available database and did not directly involve patients; it did not require approval by an ethics committee.

## Results

Brazil has faced a new COVID-19 outbreak more devastating than the beginning of the pandemic, with hospitals under increasing pressure due to the high number of hospitalizations. ICU admission, IMV requirement and in-hospital deaths also follow this trend (Figure 1). However, the macro-level does not reflect the differences in the federative units, whose territorial dimensions and demographic, socioeconomic and cultural characteristics are variable. The outbreaks desynchronization among the federative units generated atypical waves in Brazil (Figure 2), seeming to have been hit by multiple waves, unlike many countries that have showed well-defined waves. Due to its continental dimension, Brazil has failed to flatten the curves of new cases adequately.

**Figure 1.**
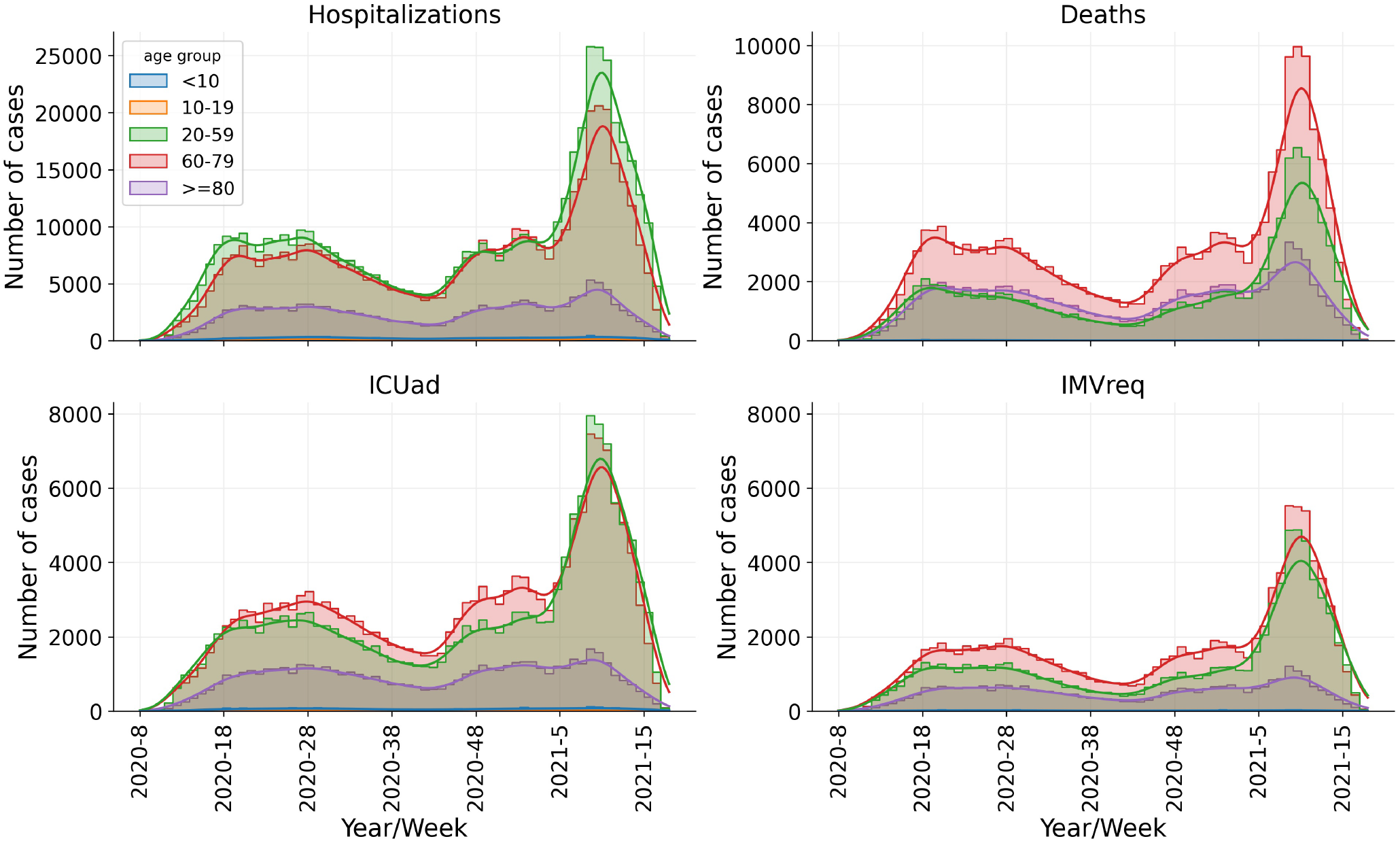
Time series of confirmed cases for hospitalizations, deaths, ICU admission and IMV requirement in Brazil. The horizontal axis (x-axis) refers to the epidemiological weeks according to the onset of symptoms. Timeline was divided into six periods of 10 weeks each, from epidemiological week 2020-8 to 2021-14.

**Figure 2.**
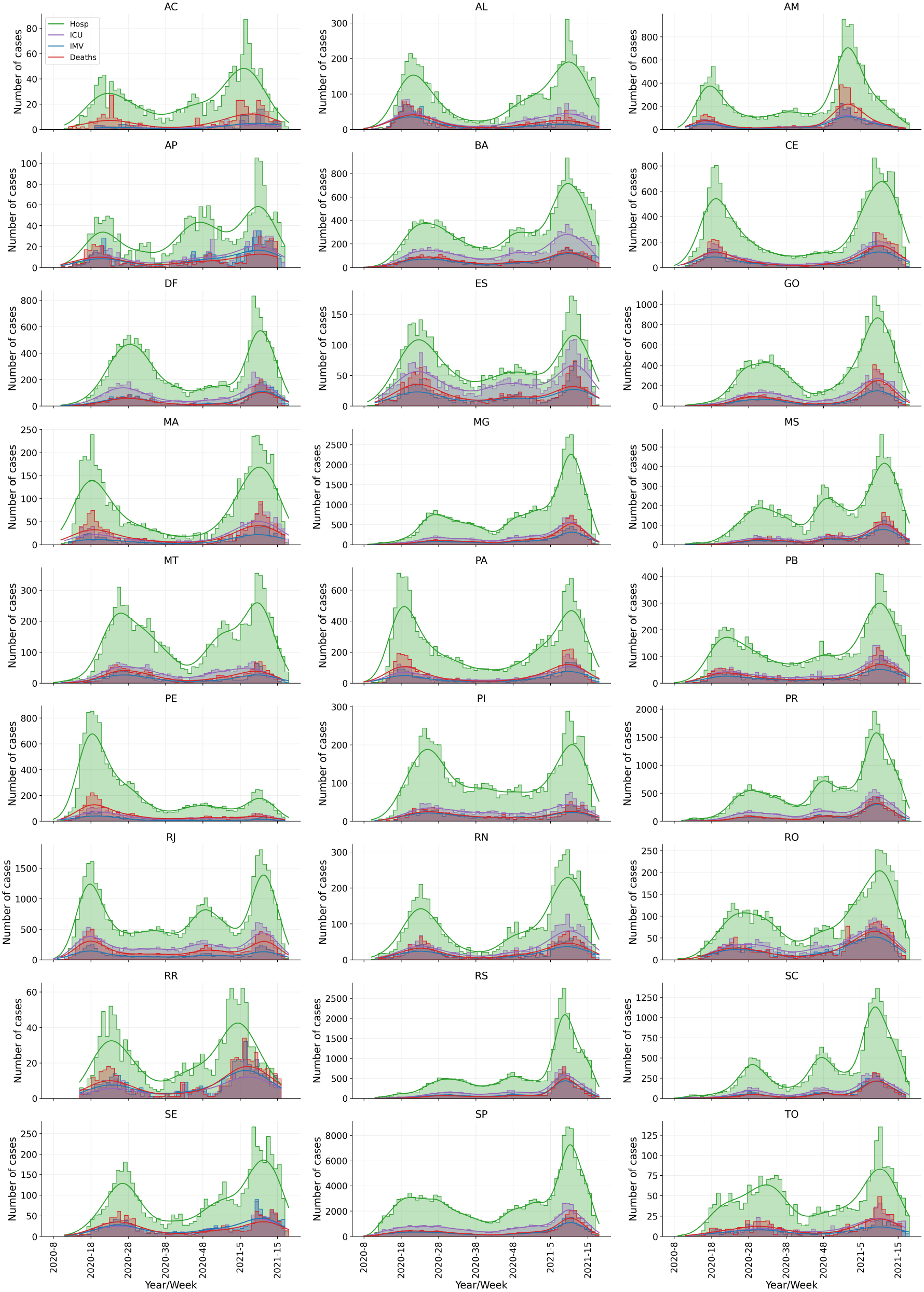
Outbreaks desynchronization (atypical waves) among the federative units in Brazil. Time series of confirmed cases for COVID-19 hospitalizations, ICU admission, IMV requirement and in-hospital deaths according to the Brazilian federative units. The horizontal axis (x-axis) refers to the epidemiological weeks according to the onset of symptoms. Timeline was divided into six periods of 10 weeks each, from epidemiological week 2020-8 to 2021-14.

Indeed, the COVID-19 pandemic in Brazil has been characterized by atypical waves resulting from subsequent outbreaks that are occurring after the initial wave, which peaked in July 2020. The beginning and the evolution pattern of the first wave varied among Brazilian regions (Figure S2), so that the initial peaks of hospitalization in each region were in AM/PA in the North, SP/RJ in the Southeast, MA/CE in the Northeast, PR in the South and MT in the Midwest (Figure 2).

In this context, we provide a spatio-temporal analysis of confirmed cases for hospitalization, ICU admission, IMV requirement and in-hospital mortality from Brazilian COVID-19 patients according to the 27 Brazilian federative units and epidemiological weeks from 2020-8 until 2021-14 (periods T1 to T6). First, the data was disaggregated into different age groups (Figure 1), because our goal is to reveal the vulnerability of young adults (patients aged 20-59 years old). Afterwards, we estimated the percentages of hospitalization, ICU admission, IMV requirement and in-hospital hCFR of COVID-19 young adults. We also calculated the risk ratios in order to compare the different periods/waves of the pandemic in Brazil.

The general analysis of the number of cases of hospitalization, ICU admission, IMV requirement, and in-hospital deaths of COVID-19 patients aged 20-59 years shows two distinct clusters (see Figure 2). The first is formed by most federative units (AC, AL, AP, BA, CE, ES, MA, MT, PA, PB, PE, PI, RJ, RN, RO, RR, SE, SP), where the case peaks occurred in T2 and T6 periods, and the second, consists of eight federative units, where the peaks were in T3 and T6 (DF, GO, MG, MS, PR, RS, SC and TO). Importantly, the Amazonas state (AM) is the only one where case peaks occurred in T1 and T5, showing the early occurrence of the second wave.

### Hospitalization

Our study revealed that hospitalized COVID-19 patients aged 20-59 years in the analyzed periods varies between 40-50% of the cases (Figure 3, Table S1). T1 accounts for the beginning of the disease evolution in the country and presents the smallest absolute number of cases. Thus, taking the T2 period as a reference (period when the first infection cases peak occurred in most federative units), we have a slight reduction of the hospitalization risk for patients aged 20-59 in: T3 (RR=0.94; 95% CI=[0.94,0.95]; *p* < 0.001), T4 (RR=0.95; 95% CI=[0.94,0.96]; *p* < 0.001), and T5 (RR=0.91; 95% CI=[0.91,0.92]; *p* < 0.001). The risk is increased in T6 (RR=1.10; 95% CI=[1.09,1.10]; *p* < 0.001), when a second wave hit Brazil.

**Figure 3.**
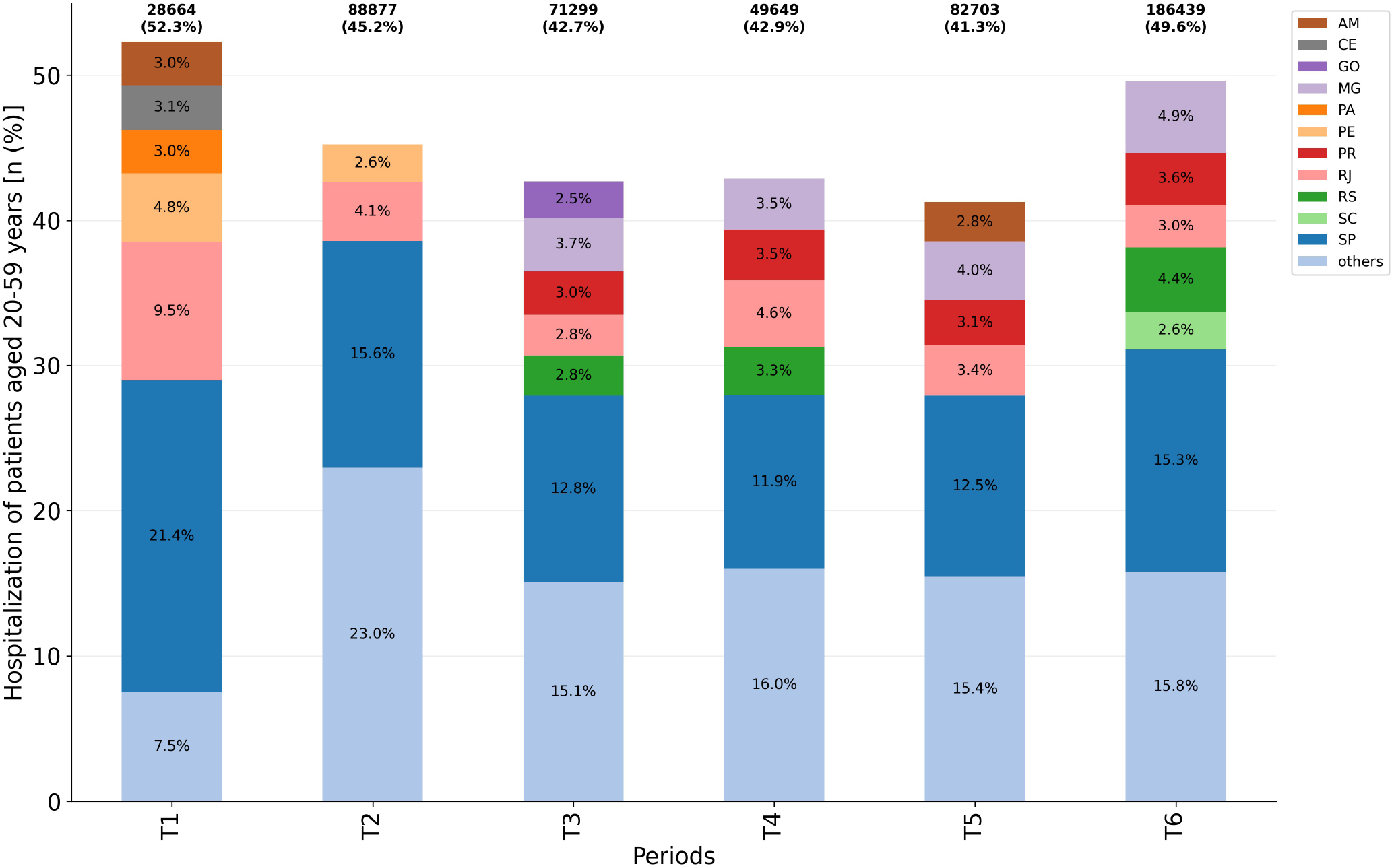
Hospitalization of patients aged 20-59 years among all age groups per considered period (*n* = 507, 631). Federative units showing proportions ≥2.5 were highlighted while ‘others’ accounts for the grouping of those presenting a proportion < 2.5.

Considering absolute numbers, it can be seen a decrease in the number of hospitalization cases in T3 (−19.8%), T4 (−44.1%) and T5 (−6.94%), compared to T2. In T6, the second wave peak, there was an increase of 109.8% in cases of hospitalization compared to the T2 period, for patients aged 20-59 years.

Among the main federative units that contributed to the high percentages of hospitalization, the highest occurrence of cases appears in the SP, varying from 11.9% to 21.4% depending on the period (Figure 3). It is important to point out that SP also has the highest population among the Brazilian federative units (see Figure S3). Therefore, over time, the hospitalization percentage did not present a high variation regarding this age group, although there has been an explosion of hospitalization cases of young adults in absolute numbers in the last analyzed period.

### Intensive Care Unit admission

ICU admission percentages for young adults aged 20-59 years did not vary significantly (≈30% −35% of hospitalized patients) over the analyzed periods (Figure 4, Table S2), with risk ratios varying from 1.02 to 1.12, when comparing periods T3, T4, T5 and T6 to T2. However, in absolute numbers there was a significant increase in the requirement, with the predominance in São Paulo, where a bigger number of hospitalizations occurred. An increase of ≈283% for ICU admission is observed in T6 compared to T4, the period with the lowest ICU admission.

**Figure 4.**
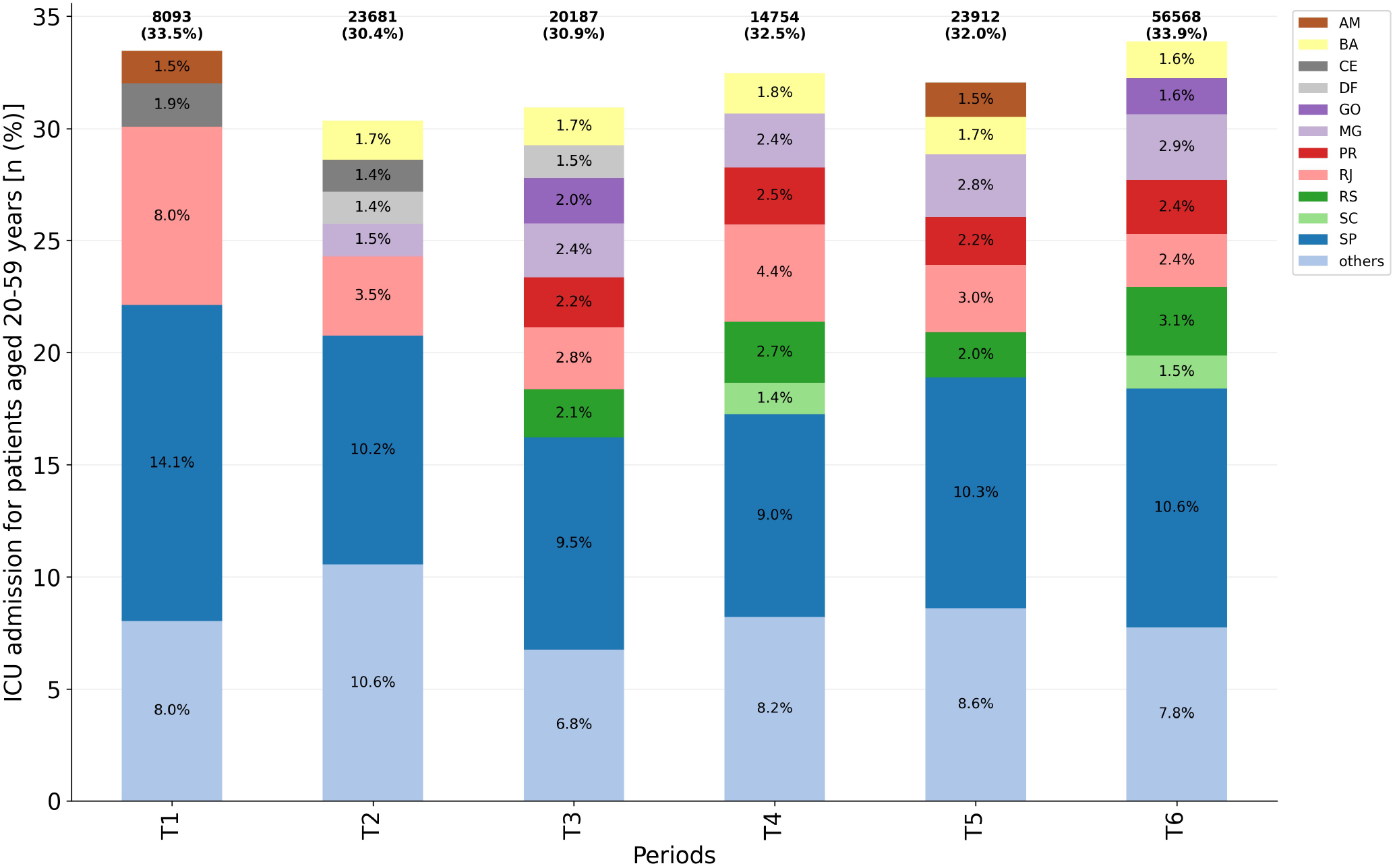
ICU admission for patients aged 20-59 years per considered period (*n* = 147, 195). Federative units showing proportions ≥ 1.4 were highlighted while ‘others’ accounts for the grouping of those presenting a proportion < 1.4.

The correlation coefficient between the number of cases of hospitalization and ICU admission in the major of federative units (23 out of 27) was ≥0.90(*p* < 0.001), demonstrating that the increase in the number of hospitalizations due to the COVID-19 can lead to the collapse of the health system, especially in localities with limited resources.

It is important to note the variation in the ICU admission proportions of different federative units, according to the periods. It can be attributed to the behaviour of desynchronized peaks among the units. For instance, Amazonas, whose case peaks occurred at T1 and T5, presented a high demand for ICU during such periods, despite its low density population (Figure S3). This shows that a larger population size is not the only factor that worsens its number of cases. Federative units of the second cluster, whose hospitalization first peak occurred later (T3), and were hit by a second wave in T6, also showed a significant increase in young adults ICU admission during those periods (DF, GO, MG, PR, RS, for instance).

### Invasive Mechanical Ventilation requirement

In contrast to ICU admission, IMV requirement varied significantly in the analyzed periods (Figure 5, Table S3), from 12.6% (T4) to 20.4% (T6). Moreover, numerically, 5,527 patients aged 20-59 years required IMV at T4, whereas in T6 this number increased to 33,506 patients. The risk ratio for IMV requirement in T6 compared to T4 was RR=1.61 (95% CI=[0.87,0.91]; *p* < 0.001).

**Figure 5.**
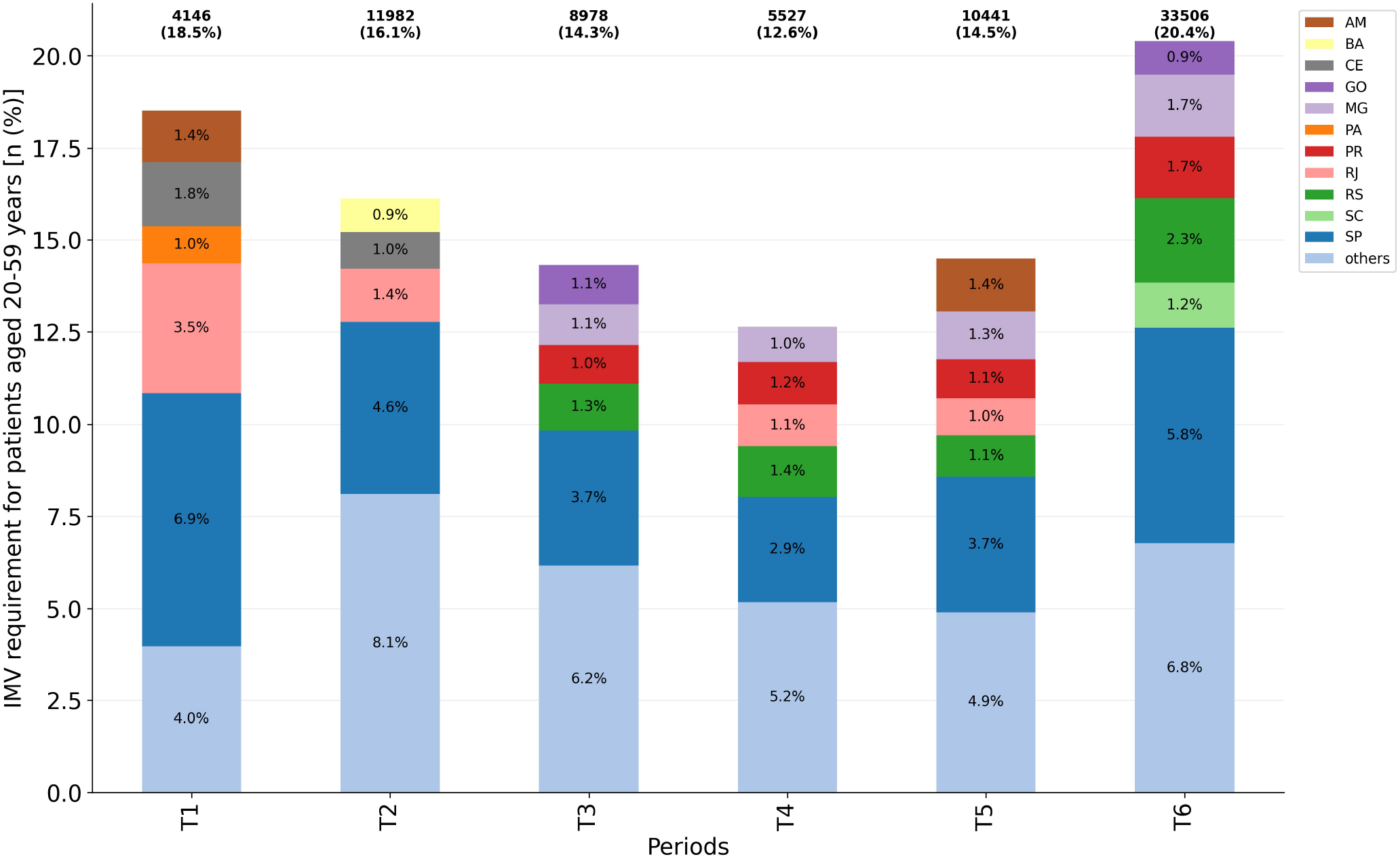
IMV requirement for patients aged 20-59 years per considered period (*n* = 74, 580). Federative units showing proportions ≥ 0.9 were highlighted while ‘others’ accounts for the grouping of those presenting a proportion < 0.9.

Taking the T2 period as a reference, we have a reduction of the IMV requirement risk for patients aged 20-59 in: T3 (RR=0.89; 95% CI=[0.87,0.91]; *p* < 0.001), T4 (RR=0.78; 95% CI=[0.76,0.81]; *p* < 0.001), and T5 (RR=0.90; 95% CI=[0.88,0.92]; *p* < 0.001). The risk is increased in T6 (RR=1.26; 95% CI=[1.24,1.29]; *p* < 0.001), during the second wave in Brazil.

In all periods, there is a predominance of cases in São Paulo, following the highest number of hospitalizations and ICU admission, as already reported. It is important to point out that different federative units contributed to the raise in IMV requirement during each period. For instance, during T1 and T2, São Paulo, Rio de Janeiro and Ceará present the main proportions, while at T6 they were São Paulo, Rio Grande do Sul, Minas Gerais and Paraná. Therefore, there is a large variation when analyzing the risk ratio across the country, according to when each place has been hit by the atypical waves in Brazil.

### In-hospital deaths

In order to estimate in-hospital deaths among hospitalized young adults, we used case fatality rate. The hCFR varied significantly among the analyzed periods (Figure 6, Table S4). The highest hCFR in T2 is given in São Paulo, Rio de Janeiro, Ceará and Pernambuco states, while at T6 accounts for São Paulo, Minas Gerais and Rio Grande do Sul, accompanying the increase in the number of hospitalization cases in these states in such periods, respectively. Amazonas state shows a significant proportion of deaths during T1 and T5.

**Figure 6.**
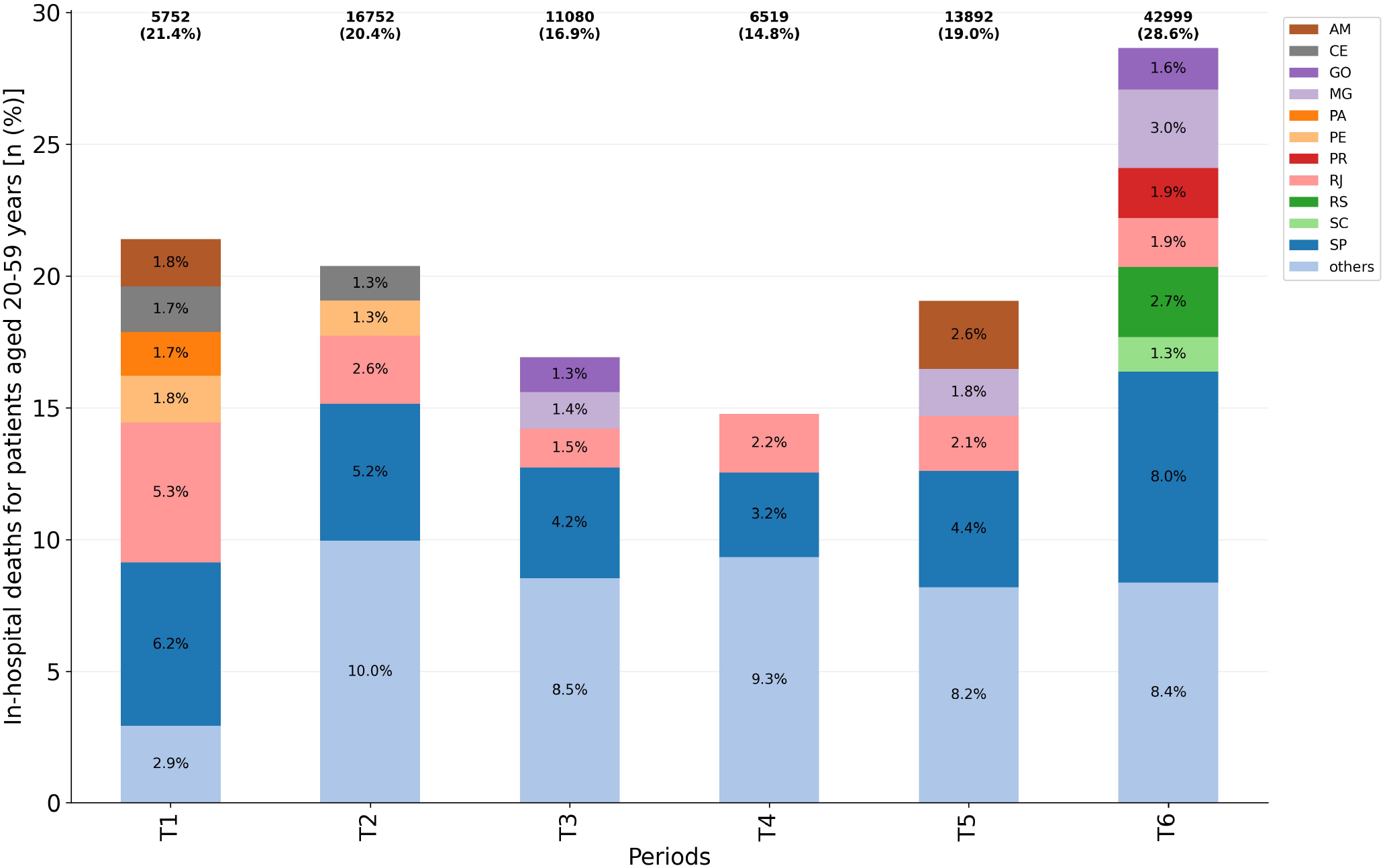
In-hospital deaths for patients aged 20-59 years with closed outcome (cure or death) per considered period (*n* = 96, 994). Federative units showing proportions ≥1.3 were highlighted while ‘others’ accounts for the grouping of those presenting a proportion < 1.3.

There was a substantial increase in mortality in absolute numbers as well as in the percentages, mainly in T5 (13,892 deaths; 19.0%) and T6 (42,999 deaths; 28.6%) compared to T4 (6,519 deaths; 14.8%). The risk ratios for such periods were RR(T5/T4)=1.29 (95% CI=[1.26,1.32]; *p* < 0.001 and RR(T6/T4)=1.94 (95% CI=[1.89,1.99]; *p* < 0.001). The lowest value (14.8%) was observed in the T4 period, which preceded the appearance of the second wave of COVID-19 in Brazil. A significant increase in the hCFR of young adults aged 20-59 years is then observed in the subsequent periods, which coincides with the variant P.1 emergence and its spread to several Brazilian states, specially in GO, MG, PR, RS and SC. Importantly, the increased hCFR in Amazonas at T5 coincides with the emergence of variant P.1 in the state. In summary, the proportions of in-hospital deaths varied among different states as well as in the different periods of the COVID-19 pandemic in Brazil, with a predominance of the São Paulo state.

### Fatality risk ratio of COVID-19 patients aged 20-59 years in the last wave vs. the first wave

Finally, we analyzed the fatality risk ratio for COVID-19 patients aged 20-59 years in the second wave relative to the first for a set of key states that showed greater prominence in previous analyzes. Table S5 summarizes the risk ratio for those states, showing an increased risk ratio for all of them. Importantly, the highest risk was observed in Rio Grande do Sul (RR=2.10; 95% CI=[1.94,2.26]; *p* < 0.001) and the lowest risks were observed in Amazonas (RR=1.16; 95% CI=[1.07,1.26]; *p* < 0.001) and Rio de Janeiro (RR=1.17; 95% CI=[1.11,1.22]; *p* < 0.001). However, these lower RRs are due to the high case fatality rate in the first outbreak (hCFR=30.94% in Amazonas; hCFR=30.04% in Rio de Janeiro), which remained high in the last outbreak (hCFR=35.99% in Amazonas; hCFR=35.05%).

### Important remarks

Overall, the least critical period of the Brazil COVID-19 pandemic for patients aged 20-59 years was in the T4 period, when there were fewer cases of hospitalization (Figure 3), IMV requirement (Figure 5) and in-hospital deaths (Figure 6), although the ICU admission (Figure 4) remained at ≈30%. Conversely, the most critical period is the last one (T6), with an explosion in the number of cases of hospitalization, deaths, and demand for ICU and IMV among young adults. Importantly, as of April 10th, 2021, the last day of analyzed data, Brazil had accumulated more than 13 million cases and approximately 345,000 deaths due to COVID-19, an increase of more than 70% since December 31st, 2020.

## Discussion

The first wave of an epidemic is due to its onset and expansion driven by community transmission, the duration of which depends on containment measures. In the first pandemic wave of COVID-19, NPIs played a significant role in flattening the curve, in the absence of pharmacological interventions such as specific drugs for treatment and vaccines. The relaxation regarding to NPIs due to returning to economic and social activities and the insufficiency in achieving herd immunity caused COVID-19 subsequent outbreaks. Several countries have faced a second wave since October 2020. In addition, the new variants emergence has affected some regions such as the UK, Brazil and South Africa.

It is important to highlight that the first outbreak in the Amazonas state started in March, 2020 and peaked in early May, 2020 and, the second, in mid-December, 2020 and extended during the month of January, 2021^22^. Our epidemiological data also indicate the main Amazonas outbreaks in those periods. Therefore, we compared T1 and T5 periods specifically for this state. The data from the other federative units were analyzed according to the two clusters we identified.

The P.1 variant was initially identified in Japan in travelers returning from Brazil in January 2021^23^. A subsequent study showed that P.1 was already circulating since December 2020 in Manaus^5^. Amazonas state health system collapsed in January 2021, with a lack of medicinal oxygen to supply the high demand of patients with COVID-19 requiring mechanical ventilation. In this emergency, several patients were transferred to other Brazilian states^24^, which may have contributed to the spread of the P.1 variant in the national territory. Variant P.1 has been identified in samples collected in January/February 2021(out and inpatients) in the Rio Grande do Sul state, and coincides with an increase in COVID-19 severe cases^25^. A preliminary study carried out with patients from the Paraná state has indicated a significant increase in case fatality rates in young and middle-aged adults in February 2021, corresponding to the spreading period of P.1 variant in the country^26^. These findings show that the variant P.1 spread rapidly across Brazilian territory and may be one of the factors responsible for the high hospitalization and mortality of younger people. Among the main states with high hCFR proportion in the T6 period, variant P.1 was detected in Minas Gerais, Paraná, Rio Grande do Sul, Santa Catarina and São Paulo states in that period^6^,27. In fact, Brazil is experiencing a numerical explosion of hospitalizations and deaths by COVID-19 in the 20-59 years age group, especially in the state of São Paulo, with a similar proportion of hospitalizations but higher proportion of deaths compared to the first wave. Unlike other countries, we had already seen a high proportion of hospitalization (47.07%) and lethality (23.0%) among young adults (20-59 years) during the first wave^11^.

Importantly, proportionally São Paulo state has contributed to the high hospitalization of young adults aged 20-59 years and with the worst percentages of ICU admission, IMV requirement and hCFR. It should be considered that São Paulo is the most populous state in Brazil with an estimated population of nearly 27 million people aged 20-59 years in 2020^28^. A recent study showed that of the 52 sequenced samples from COVID-19 patients in the São Paulo city, 84.4% were P.1 variant, that is more transmissible and infectious^27^. Also taking into account that São Paulo state has a population of ≈ 70% in working age, these factors could justify the results obtained for the age group of 20-59 years in the state, since they are numerically majority and more exposed to coronavirus due to economic needs.

In the second wave, it was expected that there would be better capacity to manage the problem with the lessons learned during the first wave and better resources to treat patients. However, with the increase in the infected people number and requiring ICU IMV resources, the mortality also increased in many countries during the second wave^29^.

The data regarding to hCFR suggest that there are one or more factors influencing the high rate in T5 and T6 periods. In this context, we can attribute this finding to several factors such as the restrictive measures reduction due to economic and social needs, relaxation regarding to personal care (wearing masks, physical distancing, and hand hygiene), the high demand for healthcare that supplanted the Brazilian system capacity, the variant P.1 emergence in various states or even the delay in seeking medical care by younger people. All of these factors, can be closely related making the reduction of hospitalization cases and deaths by COVID-19 a difficult problem to be overcome in Brazil.

Regarding the fatality risk ratio for COVID-19 patients aged 20-59 years in the second wave relative to the first wave, the risk was of more than 1.5× in Minas Gerais, Paraná, Rio Grande Sul, Santa Catarina and São Paulo (states where P.1 was detected in T6 period). Besides the P.1 variant, Amazonas was affected by a lack of medicinal oxygen, which may have led to an increase in the mortality of hospitalized young adults.

In summary, our study indicates that the atypical waves of the COVID-19 pandemic seen in Brazil are due to the desynchronization of the outbreaks that occurred in the different federal units of the country. This desynchronization is reflected in the number of cases of hospitalization, ICU admission, IMV requirement and deaths. Time series analyzes of hospitalized COVID-19 patients aged 20-59 years showed variations in hospitalization, IMV requirement and hCFR in the different periods, except for ICU admission which was ≈30% in all periods. The parameters analyzed in the different periods also showed variations among states, with São Paulo being highlighted for the highest percentages in all periods.

We acknowledge some limitations in our study. First, the SIVEP-Gripe database contains only individual information of hospitalized severe acute respiratory illness patients, not informing the daily number of infection cases in each federative unit and the respective outcomes. Therefore, we limit ourselves to analyzing data from hospitalized patients. Second, we include all patients who have been diagnosed for COVID-19 by different methods and not just by RT-PCR, since the immunological diagnosis is predominant in several federative units, due to the difficulties of carrying out tests that are more expensive. Third, we did not investigate the causes of increased hospitalization and mortality for COVID-19 patients aged 20-59 years, since this was not the main objective of the study and, in addition, there is not enough data to prove, for example, higher variant P.1 lethality.

Due to the limited vaccines availability, the vast majority of countries initially chose to vaccinate the elderly, who are more susceptible to severe COVID-19. Therefore, while there is not enough vaccine for everyone, NPIs are extremely important to contain the infection and spread of the disease among young adults, who are more exposed mainly due to work and economic needs.

At the time of preparation of this report, there is a forecast of a third wave in the Amazonas state, which serves as an alert for the entire country. Cases number stabilization at high levels, as we have seen in this second wave with ≈2, 000 deaths/day, can drive a third wave even more disastrous in Brazil and hit the younger age group more severely.

## Conclusion

Our study have shown the origin of atypical waves of the pandemic COVID-19 in aggregate data from all over Brazil, whose continental dimensions make it a country with different characteristics from the others. In addition, we have showed that the resurgence of COVID-19 outbreaks, with a high number of new cases, has significantly affected young adults in several regions of the country. A comprehensive view of the hospitalized COVID-19 patients fatality rates in the 20-59 age group, indicated significant variations in the analyzed periods and in the different federative units in Brazil.

Overall, in the period January-February 2021, there was an increase in hospitalization and death cases for young adults. Thus, our study confirms the preliminary evidence of a significant increase in cases of hospitalization, ICU admission, IMV requirement and deaths of younger COVID-19 patients.

The pandemic evolution across the country is variable due to the adoption of different public policies in Brazilian states. Prevention strategies through NPIs, especially among young adults, while there are not enough vaccines is of the extreme importance to prevent the new VOCs emergence and spread. Due to the limited availability of vaccines, the vast majority of countries initially chose to vaccinate the elderly population, who are more susceptible to severe COVID-19. However, our analysis indicate the need to focus on the younger age group, which must also be preserved, as it represents the productive force of each country, beyond the universal right of everyone to access the COVID-19 vaccine.

In summary, the study warns health managers to pay special attention to cases of COVID-19 young adult patients in states with larger populations, as the health system is at risk of collapsing.

## Data Availability

The data used in this work is publicly available.

## Author contributions

F.S.H.S., N.S.H-S. and D.L.G. conceived and designed the study. F.S.H.S., N.S.H-S., D.C.R.O., C.M.S. and D.L.G. analysed and interpreted the data. N.S.H-S., F.S.H.S. and D.L.G. wrote the manuscript. All authors reviewed earlier drafts and approved its final version.

## Additional information

### Data availability

The data used in this work is publicly available^18^.

## References

1. WHO. World Health Organization. WHO announces COVID-19 outbreak a pandemic; 2020. [Accessed May 22, 2020]. Available from: http://www.euro.who.int/en/health-topics/health-emergencies/coronavirus-covid-19/news/news/2020/3/who-announces-covid-19-outbreak-a-pandemic.

2. Davies NG, Abbott S, Barnard RC, Jarvis CI, Kucharski AJ, Munday JD, et al. Estimated transmissibility and impact of SARS-CoV-2 lineage B.1.1.7 in England. Science. 2021;372(6538). Available from: https://science.sciencemag.org/content/372/6538/eabg3055.

3. Davies NG, Jarvis CI, van Zandvoort K, Clifford S, Sun FY, Funk S, et al. Increased mortality in community-tested cases of SARS-CoV-2 lineage B.1.1.7. Nature. 2021 May;593(7858):270–274. Available from: https://doi.org/10.1038/s41586-021-03426-1.

4. Coutinho RM, Marquitti FMD, Ferreira LS, Borges ME, da Silva RLP, Canton O, et al. Model-based estimation of transmissibility and reinfection of SARS-CoV-2 P.1 variant. medRxiv. 2021. Available from: https://www.medrxiv.org/content/early/2021/03/09/2021.03.03.21252706.

5. Naveca F, Costa C. Caracterização genética do SARS-CoV-2 circulante no Estado do Amazonas. Nota Técnica Conjunta Nº 09/Fiocruz/ILMD E FVS-AM; 2021. Published online Jan 28. [Accessed March 15, 2021]. Available from: https://amazonia.fiocruz.br/wp-content/uploads/2021/01/NOTA-TE%CC%81CNICA-CONJUNTA-N%C2%BA-09.2021.FVS-AM-X-ILMD.FICRUZ-AM-28.01.2021.pdf.

6. Observatório Covid-19 Fiocruz. Fiocruz detecta mutação associada a variantes de preocupação do Sars-Cov-2 em diversos estados do país. Observatório Covid-19 Fiocruz: Informação para ação; 2021. Published online March, 4. [Accessed March 15, 2021]. Available from: https://portal.fiocruz.br/sites/portal.fiocruz.br/files/documentos/comunicado_variantes_de_preocupacao_fiocruz_2_2021-03-04.pdf.

7. Brasil. Instituto Brasileiro de Geografia e Estatística (IBGE). Brasil em síntese: território; 2021. [Accessed March 17, 2021]. Available from: https://brasilemsintese.ibge.gov.br/territorio.html.

8. Castro MC, Massuda A, Almeida G, Menezes-Filho NA, Andrade MV, de Souza Noronha KVM, et al. Brazil’s unified health system: the first 30 years and prospects for the future. The Lancet. 2019 Jul;394(10195):345–356. Available from: https://doi.org/10.1016/S0140-6736(19)31243-7.

9. Rocha R, Atun R, Massuda A, Rache B, Spinola P, Nunes L, et al. Effect of socioeconomic inequalities and vulnerabilities on health-system preparedness and response to COVID-19 in Brazil: a comprehensive analysis. The Lancet Global Health. 2021 May. Available from: https://doi.org/10.1016/S2214-109X(21)00081-4.

10. Baqui P, Bica I, Marra V, Ercole A, van der Schaar M. Ethnic and regional variations in hospital mortality from COVID-19 in Brazil: a cross-sectional observational study. The Lancet Global Health. 2020 Aug;8(8):e1018–e1026. Available from: https://doi.org/10.1016/S2214-109X(20)30285-0.

11. de Souza FSH, Hojo-Souza NS, Batista BDdO, da Silva CM, Guidoni DL. On the analysis of mortality risk factors for hospitalized COVID-19 patients: A data-driven study using the major Brazilian database. PLOS ONE. 2021 03;16(3):1–21. Available from: https://doi.org/10.1371/journal.pone.0248580.

12. Fu L, Wang B, Yuan T, Chen X, Ao Y, Fitzpatrick T, et al. Clinical characteristics of coronavirus disease 2019 (COVID-19) in China: A systematic review and meta-analysis. Journal of Infection. 2020;80(6):656–665. Available from: https://doi.org/10.1016/j.jinf.2020.03.041.

13. Kim L, Garg S, O’Halloran A, Whitaker M, Pham H, Anderson EJ, et al. Risk Factors for Intensive Care Unit Admission and In-hospital Mortality among Hospitalized Adults Identified through the U.S. Coronavirus Disease 2019 (COVID-19)-Associated Hospitalization Surveillance Network (COVID-NET).Clinical Infectious Diseases. 2020 07. Ciaa1012. Available from: https://doi.org/10.1093/cid/ciaa1012.

14. Petrilli CM, Jones SA, Yang J, Rajagopalan H, O’Donnell L, Chernyak Y, et al. Factors associated with hospital admission and critical illness among 5279 people with coronavirus disease 2019 in New York City: prospective cohort study. BMJ. 2020;369:m1966. Available from: https://www.bmj.com/content/369/bmj.m1966.

15. Brasil. Instituto Brasileiro de Geografia e Estatística (IBGE). População em idade ativa (PIA); 2021. [Accessed March 17, 2021]. Available from: https://www.ibge.gov.br/apps/populacao/projecao//index.html.

16. Boletim do Observatório Covid-19: Informação para ação. Semanas epidemiológicas 10 e 11. Observatório Covid-19 Fiocruz: Informação para ação; 2021. Published online March, 4. [Accessed May 17, 2021]. Available from: https://portal.fiocruz.br/sites/portal.fiocruz.br/files/documentos/boletim_covid_2021-semanas_10-11-red.pdf.

17. de Souza FSH, Hojo-Souza NS, da Silva CM, Guidoni DL. Second wave of COVID-19 in Brazil: younger at higher risk. European Journal of Epidemiology. 2021 Apr;36(4):441–443. Available from: https://doi.org/10.1007/s10654-021-00750-8.

18. Brasil. Ministério da Saúde. Sistema de Informação de Vigilância Epidemiológica da Gripe, SIVEP-Gripe; 2021. [Accessed March 17, 2021]. Available from: https://opendatasus.saude.gov.br/.

19. Ministério da Saúde. COVID-19 Vacinação, Doses Aplicadas.; 2021. [Accessed May 21, 2021]. Available from: https://qsprod.saude.gov.br/extensions/DEMAS_C19Vacina/DEMAS_C19Vacina.html.

20. Siegel S, Castellan NJ. Nonparametric statistics for the behavioral sciences. 2nd ed. McGraw–Hill, Inc.; 1988.

21. Hosmer DW, Lemeshow S. Applied logistic regression. John Wiley and Sons; 2000.

22. Naveca FG, Nascimento V, de Souza VC, Corado AdL, Nascimento F, Silva G, et al. COVID-19 in Amazonas, Brazil, was driven by the persistence of endemic lineages and P.1 emergence. Nature Medicine. 2021 May. Available from: https://doi.org/10.1038/s41591-021-01378-7.

23. Fujino T, Nomoto H, Kutsuna S, Ujiie M, Suzuki T, Sato R, et al. Novel SARS-CoV-2 Variant in Travelers from Brazil to Japan. Emerging Infectious Diseases. 2021 Apr;27(4). Available from: https://doi.org/10.3201/eid2704.210138.

24. AEROFLAP. Notícias. Forças Armadas transportaram 291 pacientes do Amazonas para outros estados.; 2021. [Accessed April 12, 2021]. Available from: https://www.aeroflap.com.br/forcas-armadas-transportaram-291-pacientes-do-amazonas-para-outros-estados/.

25. Martins AF, Zavascki AP, Wink PL, Volpato FCZ, Monteiro FL, Rosset C, et al. Detection of SARS-CoV-2 lineage P.1 in patients from a region with exponentially increasing hospitalisation rate, February 2021, Rio Grande do Sul, Southern Brazil. Eurosurveillance. 2021;26(12). Available from: https://www.eurosurveillance.org/content/10.2807/1560-7917.ES.2021.26.12.2100276.

26. de Oliveira MHS, Lippi G, Henry BM. Sudden rise in COVID-19 case fatality among young and middle-aged adults in the south of Brazil after identification of the novel B.1.1.28.1 (P.1) SARS-CoV-2 strain: analysis of data from the state of Parana. medRxiv. 2021. Available from: https://www.medrxiv.org/content/early/2021/03/26/2021.03.24.21254046.

27. Barbosa GR, Moreira LVL, Justo AFO, Perosa AH, de Souza Luna LK, Chaves APC, et al. Rapid spread and high impact of the variant of concern P.1 in the largest city of Brazil. Journal of Infection. 2021 Apr. Available from: https://doi.org/10.1016/j.jinf.2021.04.008.

28. Brasil. Instituto Brasileiro de Geografia e Estatística (IBGE). Projeções e estimativas da população do Brasil e das Unidades da Federação.; 2021. [Accessed March 17, 2021]. Available from: https://www.ibge.gov.br/estatisticas/sociais/populacao/9109-projecao-da-populacao.html?=&t=resultados.

29. Graichen H. What is the difference between the first and the second/third wave of Covid-19? – German perspective. Journal of Orthopaedics. 2021;24:A1–A3. Available from: https://doi.org/10.1016/j.jor.2021.01.011.

