## Supplementary Material for "Young adults vulnerability to COVID-19 in Brazil: an overview across the country"

### ABSTRACT

Brazil is a country of continental dimensions, where many smaller countries would fit. In addition to demographic, socioeconomic, and cultural differences, hospital infrastructure and healthcare varies across all 27 federative units. Therefore, the evolution of COVID-19 pandemic did not manifest itself in a homogeneous and predictable trend across the nation. In late 2020 and early 2021, new waves of the COVID-19 outbreak have caused an unprecedented sanitary collapse in Brazil. Unlike the first COVID-19 wave, in subsequent waves, preliminary evidence has pointed to an increase in the daily reported cases among younger people being hospitalized, overloading the healthcare system. In this comprehensive retrospective study, confirmed cases of hospitalization, ICU admission, IMV requirement and in-hospital death from Brazilian COVID-19 patients throughout 2020 until the beginning of 2021 were analyzed through a spatio-temporal study for patients aged 20-59 years. All Brazilian federative units had their data disaggregated in six periods of ten epidemiological weeks each. We found that there is a wide variation in the waves dynamic due to SARS-CoV-2 infection, both in the first and in subsequent outbreaks in different federative units over the analyzed periods. As a result, atypical waves can be seen in the Brazil data as a whole. The analysis showed that Brazil is experiencing a numerical explosion of hospitalizations and deaths for patients aged 20-59 years, especially in the state of São Paulo, with a similar proportion of hospitalizations for this age group but higher proportion of deaths compared to the first wave.

### Supplementary Material

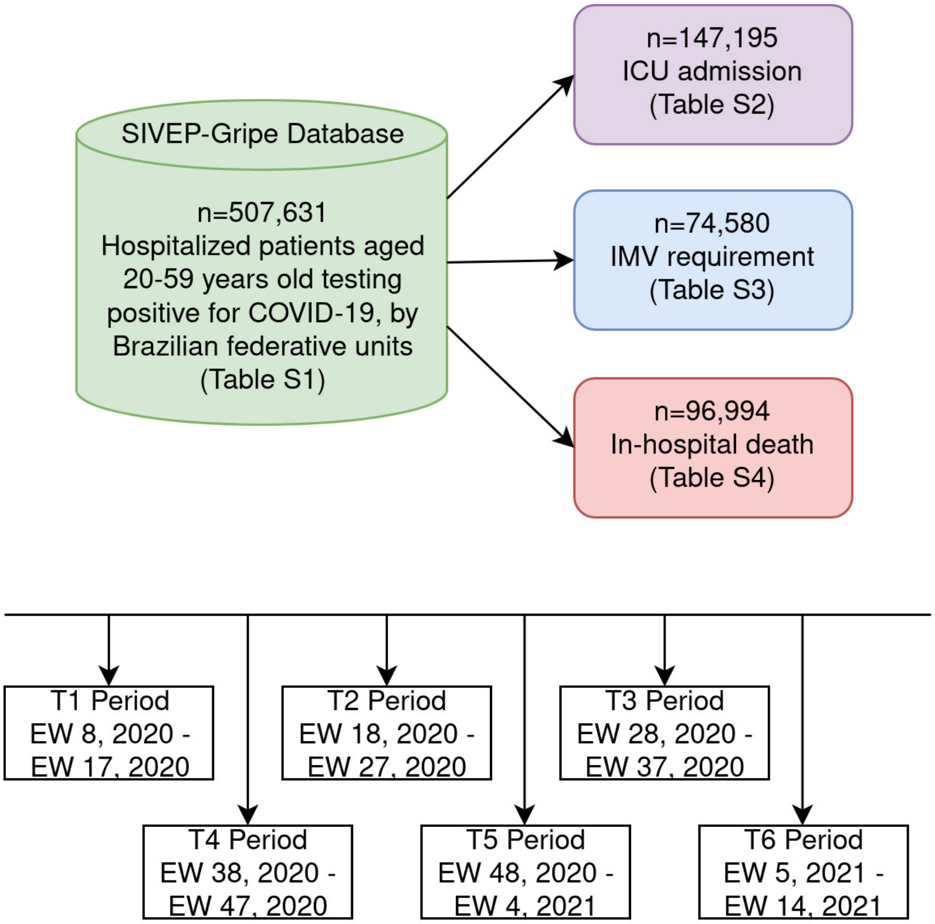


**Figure S1. Diagram of the study population.** Data regarding hospitalized patients aged 20-59 years old testing positive for COVID-19 retrieved from SIVEP-Gripe Database and stratified to account for ICU admission, IMV requirement and in-hospital death. Six periods of 10 weeks each, according to epidemiological weeks of the onset of symptoms.

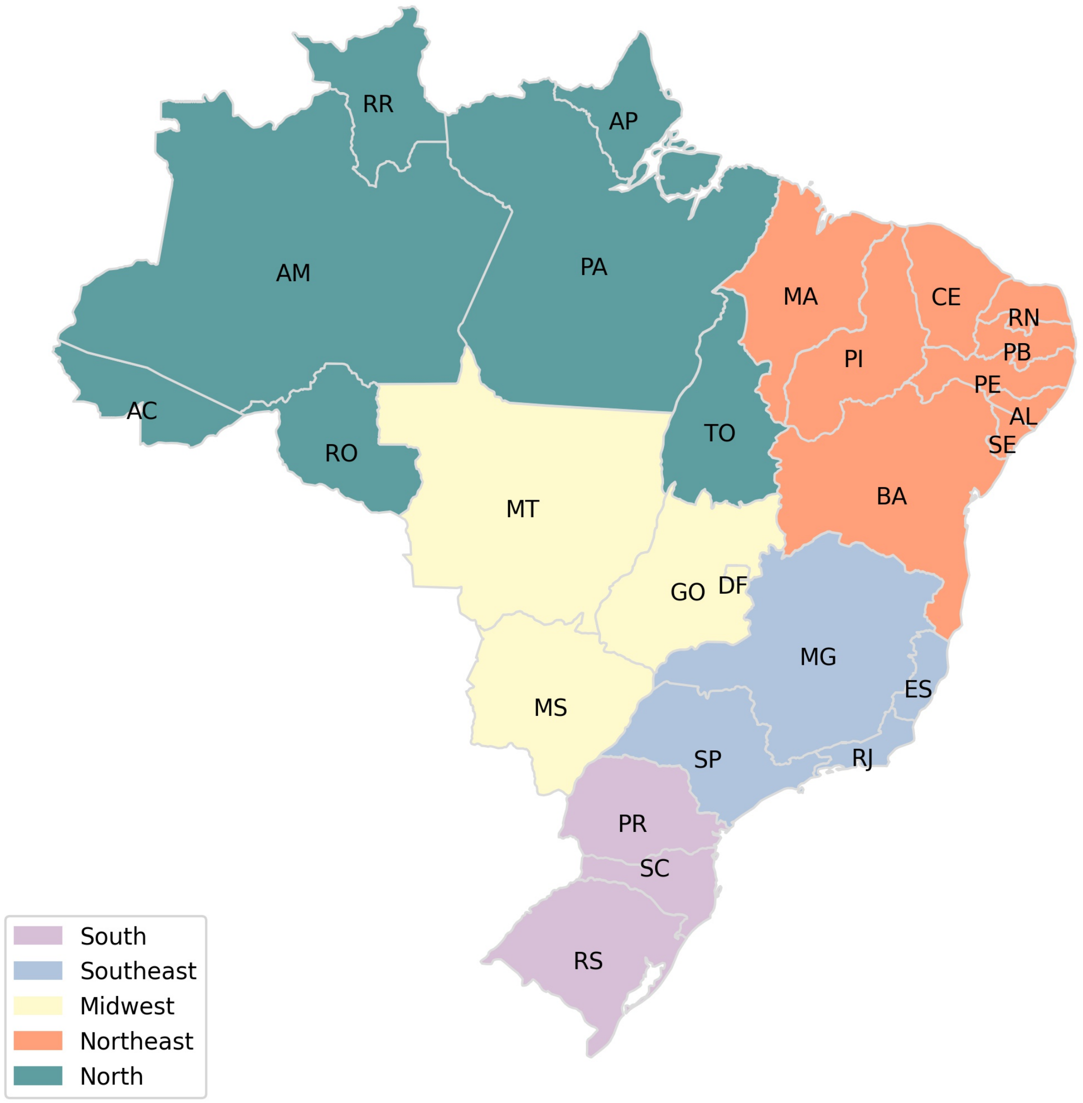


**Figure S2. Brazilian Regions**. Brazil is divided into five regions by the Brazilian Institute of Geography and Statistics (IBGE). The federative units are grouped according to geographic, social and economic factors. Brazilian federative units: Acre (AC), Alagoas (AL), Amazonas (AM), Amapá (AP), Bahia (BA), Ceará (CE), Distrito Federal (DF), Espírito Santo (ES), Goiás (GO), Maranhão (MA), Minas Gerais (MG), Mato Grosso do Sul (MS), Mato Grosso (MT), Pará (PA), Paraíba (PB), Pernambuco (PE), Piauí (PI), Paraná (PR), Rio de Janeiro (RJ), Rio Grande do Norte (RN), Rondônia (RO), Roraima (RR), Rio Grande do Sul (RS), Santa Catarina (SC), Sergipe (SE), São Paulo (SP), Tocantins (TO).

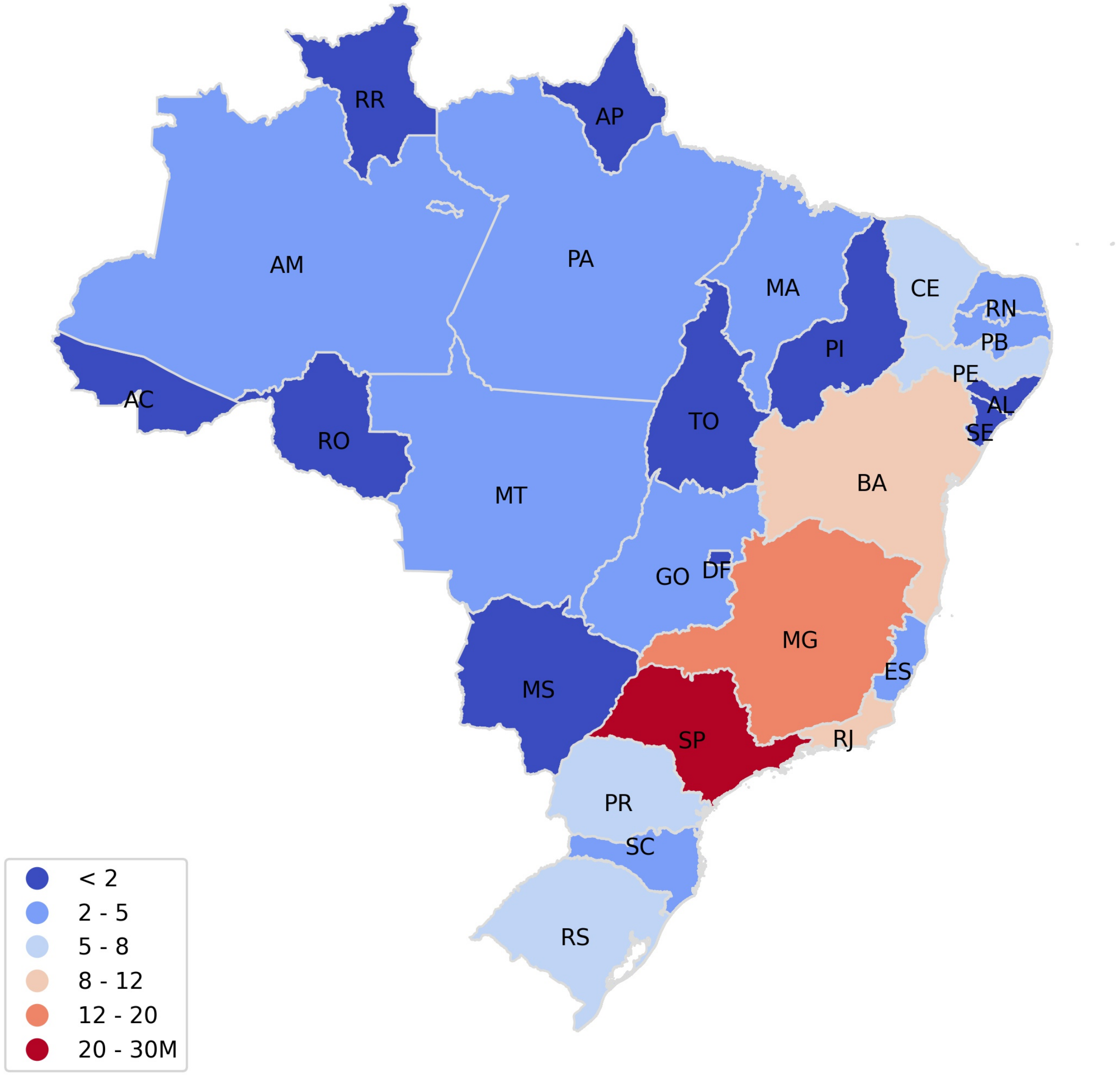


**Figure S3. Population aged 20-59 in federative units of Brazil**. Estimates for the total population aged 20-59 according to the Brazilian Institute of Geography and Statistics (IBGE). Numbers are given in millions.

**Table S1.** Number of SARI hospitalizations among COVID-19 patients according to the Brazilian federative unit, age group and outbreak periods.

| | T1 | (%) | T2 | (%) | T3 | (%) | T4 | (%) | T5 | (%) | T6 | (%) | Total | (%) |
|---|---|---|---|---|---|---|---|---|---|---|---|---|---|---|
| **all** | **54,810** | **(100.00)** | **196,510** | **(100.00)** | **166,982** | **(100.00)** | **115,762** | **(100.00)** | **200,450** | **(100.00)** | **375,941** | **(100.00)** | **1,110,455** | **(100.00)** |
| **20-59** | **28,664** | **(52.30)** | **88,877** | **(45.23)** | **71,299** | **(42.70)** | **49,649** | **(42.89)** | **82,703** | **(41.26)** | **186,439** | **(49.59)** | **507,631** | **(45.71)** |
| AC | 45 | (0.08) | 308 | (0.16) | 143 | (0.09) | 171 | (0.15) | 282 | (0.14) | 444 | (0.12) | 1,393 | (0.13) |
| AL | 216 | (0.39) | 1,563 | (0.80) | 373 | (0.22) | 283 | (0.24) | 887 | (0.44) | 1,901 | (0.51) | 5,223 | (0.47) |
| AM | 1,649 | (3.01) | 2,382 | (1.21) | 1,163 | (0.70) | 1,424 | (1.23) | 5,533 | (2.76) | 2,785 | (0.74) | 14,936 | (1.35) |
| AP | 56 | (0.10) | 341 | (0.17) | 136 | (0.08) | 339 | (0.29) | 347 | (0.17) | 598 | (0.16) | 1,817 | (0.16) |
| BA | 374 | (0.68) | 3,412 | (1.74) | 2,600 | (1.56) | 1,805 | (1.56) | 2,982 | (1.49) | 6,674 | (1.78) | 17,847 | (1.61) |
| CE | 1,673 | (3.05) | 4,672 | (2.38) | 1,392 | (0.83) | 959 | (0.83) | 1,613 | (0.80) | 6,758 | (1.80) | 17,067 | (1.54) |
| DF | 185 | (0.34) | 2,930 | (1.49) | 3,823 | (2.29) | 1,335 | (1.15) | 1,467 | (0.73) | 4,979 | (1.32) | 14,719 | (1.33) |
| ES | 215 | (0.39) | 1,153 | (0.59) | 508 | (0.30) | 468 | (0.40) | 526 | (0.26) | 1,127 | (0.30) | 3,997 | (0.36) |
| GO | 131 | (0.24) | 1,940 | (0.99) | 4,203 | (2.52) | 1,836 | (1.59) | 2,476 | (1.24) | 7,999 | (2.13) | 18,585 | (1.67) |
| MA | 717 | (1.31) | 1,130 | (0.58) | 374 | (0.22) | 179 | (0.15) | 430 | (0.21) | 1,855 | (0.49) | 4,685 | (0.42) |
| MG | 367 | (0.67) | 4,556 | (2.32) | 6,229 | (3.73) | 4,080 | (3.52) | 8,061 | (4.02) | 18,507 | (4.92) | 41,800 | (3.76) |
| MS | 49 | (0.09) | 699 | (0.36) | 1,823 | (1.09) | 1,282 | (1.11) | 2,079 | (1.04) | 3,631 | (0.97) | 9,563 | (0.86) |
| MT | 95 | (0.17) | 1,767 | (0.90) | 1,661 | (0.99) | 648 | (0.56) | 1,465 | (0.73) | 2,423 | (0.64) | 8,059 | (0.73) |
| PA | 1,643 | (3.00) | 3,976 | (2.02) | 1,440 | (0.86) | 862 | (0.74) | 1,829 | (0.91) | 4,479 | (1.19) | 14,229 | (1.28) |
| PB | 215 | (0.39) | 1,720 | (0.88) | 964 | (0.58) | 830 | (0.72) | 1,028 | (0.51) | 2,872 | (0.76) | 7,629 | (0.69) |
| PE | 2,611 | (4.76) | 5,070 | (2.58) | 1,685 | (1.01) | 963 | (0.83) | 1,063 | (0.53) | 1,618 | (0.43) | 13,010 | (1.17) |
| PI | 134 | (0.24) | 1,716 | (0.87) | 1,108 | (0.66) | 839 | (0.72) | 779 | (0.39) | 1,968 | (0.52) | 6,544 | (0.59) |
| PR | 284 | (0.52) | 2,922 | (1.49) | 4,990 | (2.99) | 4,078 | (3.52) | 6,276 | (3.13) | 13,410 | (3.57) | 31,960 | (2.88) |
| RJ | 5,227 | (9.54) | 8,086 | (4.11) | 4,693 | (2.81) | 5,334 | (4.61) | 6,913 | (3.45) | 11,314 | (3.01) | 41,567 | (3.74) |
| RN | 158 | (0.29) | 1,420 | (0.72) | 461 | (0.28) | 326 | (0.28) | 817 | (0.41) | 2,352 | (0.63) | 5,534 | (0.50) |
| RO | 87 | (0.16) | 882 | (0.45) | 912 | (0.55) | 441 | (0.38) | 894 | (0.45) | 1,987 | (0.53) | 5,203 | (0.47) |
| RR | 22 | (0.04) | 347 | (0.18) | 125 | (0.07) | 147 | (0.13) | 331 | (0.17) | 314 | (0.08) | 1,286 | (0.12) |
| RS | 440 | (0.80) | 2,304 | (1.17) | 4,596 | (2.75) | 3,833 | (3.31) | 4,807 | (2.40) | 16,614 | (4.42) | 32,594 | (2.94) |
| SC | 246 | (0.45) | 1,545 | (0.79) | 3,086 | (1.85) | 2,771 | (2.39) | 3,660 | (1.83) | 9,704 | (2.58) | 21,012 | (1.89) |
| SE | 56 | (0.10) | 1,017 | (0.52) | 728 | (0.44) | 370 | (0.32) | 876 | (0.44) | 1,771 | (0.47) | 4,818 | (0.43) |
| SP | 11,740 | (21.42) | 30,605 | (15.57) | 21,437 | (12.84) | 13,811 | (11.93) | 24,966 | (12.45) | 57,544 | (15.31) | 160,103 | (14.42) |
| TO | 29 | (0.05) | 414 | (0.21) | 646 | (0.39) | 235 | (0.20) | 316 | (0.16) | 811 | (0.22) | 2,451 | (0.22) |
| **other ages** | **26,146** | **(47.70)** | **107,633** | **(54.77)** | **95,683** | **(57.30)** | **66,113** | **(57.11)** | **117,747** | **(58.74)** | **189,502** | **(50.41)** | **602,824** | **(54.29)** |
| AC | 25 | (0.05) | 319 | (0.16) | 184 | (0.11) | 118 | (0.10) | 237 | (0.12) | 442 | (0.12) | 1,325 | (0.12) |
| AL | 212 | (0.39) | 2,271 | (1.16) | 648 | (0.39) | 354 | (0.31) | 1,188 | (0.59) | 1,881 | (0.50) | 6,554 | (0.59) |
| AM | 1,814 | (3.31) | 2,903 | (1.48) | 1,363 | (0.82) | 1,649 | (1.42) | 5,440 | (2.71) | 2,532 | (0.67) | 15,701 | (1.41) |
| AP | 42 | (0.08) | 404 | (0.21) | 211 | (0.13) | 308 | (0.27) | 402 | (0.20) | 434 | (0.12) | 1,801 | (0.16) |
| BA | 334 | (0.61) | 4,583 | (2.33) | 4,163 | (2.49) | 2,707 | (2.34) | 4,588 | (2.29) | 7,350 | (1.96) | 23,725 | (2.14) |
| CE | 2,047 | (3.73) | 7,980 | (4.06) | 2,598 | (1.56) | 1,612 | (1.39) | 2,303 | (1.15) | 7,734 | (2.06) | 24,274 | (2.19) |
| DF | 107 | (0.20) | 2,540 | (1.29) | 4,156 | (2.49) | 1,584 | (1.37) | 1,633 | (0.81) | 4,078 | (1.08) | 14,098 | (1.27) |
| ES | 238 | (0.43) | 1,906 | (0.97) | 874 | (0.52) | 739 | (0.64) | 952 | (0.47) | 1,354 | (0.36) | 6,063 | (0.55) |
| GO | 102 | (0.19) | 2,042 | (1.04) | 5,554 | (3.33) | 2,599 | (2.25) | 2,877 | (1.44) | 6,942 | (1.85) | 20,116 | (1.81) |
| MA | 721 | (1.32) | 2,057 | (1.05) | 642 | (0.38) | 298 | (0.26) | 527 | (0.26) | 2,238 | (0.60) | 6,483 | (0.58) |
| MG | 368 | (0.67) | 5,351 | (2.72) | 9,845 | (5.90) | 6,457 | (5.58) | 14,390 | (7.18) | 22,866 | (6.08) | 59,277 | (5.34) |
| MS | 27 | (0.05) | 481 | (0.24) | 1,924 | (1.15) | 1,229 | (1.06) | 2,270 | (1.13) | 2,846 | (0.76) | 8,777 | (0.79) |
| MT | 40 | (0.07) | 1,201 | (0.61) | 1,658 | (0.99) | 763 | (0.66) | 1,395 | (0.70) | 1,664 | (0.44) | 6,721 | (0.61) |
| PA | 1,536 | (2.80) | 5,237 | (2.67) | 1,850 | (1.11) | 1,157 | (1.00) | 2,449 | (1.22) | 5,247 | (1.40) | 17,476 | (1.57) |
| PB | 199 | (0.36) | 2,539 | (1.29) | 1,589 | (0.95) | 996 | (0.86) | 1,665 | (0.83) | 2,962 | (0.79) | 9,950 | (0.90) |
| PE | 2,144 | (3.91) | 6,999 | (3.56) | 2,757 | (1.65) | 1,397 | (1.21) | 1,859 | (0.93) | 1,884 | (0.50) | 17,040 | (1.53) |
| PI | 109 | (0.20) | 2,136 | (1.09) | 1,791 | (1.07) | 1,089 | (0.94) | 1,000 | (0.50) | 2,251 | (0.60) | 8,376 | (0.75) |
| PR | 240 | (0.44) | 2,701 | (1.37) | 5,515 | (3.30) | 4,719 | (4.08) | 8,366 | (4.17) | 12,365 | (3.29) | 33,906 | (3.05) |
| RJ | 5,659 | (10.32) | 12,912 | (6.57) | 7,619 | (4.56) | 8,942 | (7.72) | 14,226 | (7.10) | 13,944 | (3.71) | 63,302 | (5.70) |
| RN | 130 | (0.24) | 2,030 | (1.03) | 795 | (0.48) | 427 | (0.37) | 1,172 | (0.58) | 2,359 | (0.63) | 6,913 | (0.62) |
| RO | 67 | (0.12) | 804 | (0.41) | 833 | (0.50) | 427 | (0.37) | 1,005 | (0.50) | 1,547 | (0.41) | 4,683 | (0.42) |
| RR | 17 | (0.03) | 408 | (0.21) | 153 | (0.09) | 120 | (0.10) | 297 | (0.15) | 259 | (0.07) | 1,254 | (0.11) |
| RS | 348 | (0.63) | 2,400 | (1.22) | 6,597 | (3.95) | 5,508 | (4.76) | 8,165 | (4.07) | 18,209 | (4.84) | 41,227 | (3.71) |
| SC | 179 | (0.33) | 1,332 | (0.68) | 3,549 | (2.13) | 3,357 | (2.90) | 5,249 | (2.62) | 9,174 | (2.44) | 22,840 | (2.06) |
| SE | 54 | (0.10) | 1,533 | (0.78) | 1,177 | (0.70) | 642 | (0.55) | 1,359 | (0.68) | 1,857 | (0.49) | 6,622 | (0.60) |
| SP | 9,368 | (17.09) | 32,144 | (16.36) | 26,748 | (16.02) | 16,575 | (14.32) | 32,316 | (16.12) | 54,121 | (14.40) | 171,272 | (15.42) |
| TO | 19 | (0.03) | 420 | (0.21) | 890 | (0.53) | 340 | (0.29) | 417 | (0.21) | 962 | (0.26) | 3,048 | (0.27) |

**Table S2.** Number of ICU admissions among COVID-19 patients aged 20-59 according to the Brazilian federative unit and outbreak periods. **'all'** accounts for hospitalized patients owning ICU information.

| | T1 | (%) | T2 | (%) | T3 | (%) | T4 | (%) | T5 | (%) | T6 | (%) | Total | (%) |
|---|---|---|---|---|---|---|---|---|---|---|---|---|---|---|
| **all** | **24,181** | **(100.00)** | **78,023** | **(100.00)** | **65,261** | **(100.00)** | **45,430** | **(100.00)** | **74,628** | **(100.00)** | **166,956** | **(100.00)** | **454,479** | **(100.00)** |
| **ICU** | **8,093** | **(33.47)** | **23,681** | **(30.35)** | **20,187** | **(30.93)** | **14,754** | **(32.48)** | **23,912** | **(32.04)** | **56,568** | **(33.88)** | **147,195** | **(32.39)** |
| AC | 0 | (0.00) | 12 | (0.02) | 11 | (0.02) | 5 | (0.01) | 19 | (0.03) | 36 | (0.02) | 83 | (0.02) |
| AL | 87 | (0.36) | 510 | (0.65) | 108 | (0.17) | 86 | (0.19) | 264 | (0.35) | 488 | (0.29) | 1,543 | (0.34) |
| AM | 353 | (1.46) | 289 | (0.37) | 163 | (0.25) | 242 | (0.53) | 1,135 | (1.52) | 561 | (0.34) | 2,743 | (0.60) |
| AP | 16 | (0.07) | 121 | (0.16) | 34 | (0.05) | 65 | (0.14) | 95 | (0.13) | 246 | (0.15) | 577 | (0.13) |
| BA | 168 | (0.69) | 1,358 | (1.74) | 1,089 | (1.67) | 819 | (1.80) | 1,243 | (1.67) | 2,735 | (1.64) | 7,412 | (1.63) |
| CE | 464 | (1.92) | 1,104 | (1.41) | 387 | (0.59) | 260 | (0.57) | 504 | (0.68) | 1,890 | (1.13) | 4,609 | (1.01) |
| DF | 76 | (0.31) | 1,114 | (1.43) | 954 | (1.46) | 345 | (0.76) | 510 | (0.68) | 1,596 | (0.96) | 4,595 | (1.01) |
| ES | 151 | (0.62) | 615 | (0.79) | 273 | (0.42) | 329 | (0.72) | 332 | (0.44) | 676 | (0.40) | 2,376 | (0.52) |
| GO | 55 | (0.23) | 644 | (0.83) | 1,320 | (2.02) | 614 | (1.35) | 897 | (1.20) | 2,664 | (1.60) | 6,194 | (1.36) |
| MA | 126 | (0.52) | 195 | (0.25) | 93 | (0.14) | 62 | (0.14) | 131 | (0.18) | 593 | (0.36) | 1,200 | (0.26) |
| MG | 128 | (0.53) | 1,163 | (1.49) | 1,594 | (2.44) | 1,091 | (2.40) | 2,089 | (2.80) | 4,903 | (2.94) | 10,968 | (2.41) |
| MS | 13 | (0.05) | 127 | (0.16) | 339 | (0.52) | 238 | (0.52) | 410 | (0.55) | 895 | (0.54) | 2,022 | (0.44) |
| MT | 29 | (0.12) | 440 | (0.56) | 449 | (0.69) | 176 | (0.39) | 370 | (0.50) | 459 | (0.27) | 1,923 | (0.42) |
| PA | 306 | (1.27) | 635 | (0.81) | 238 | (0.36) | 191 | (0.42) | 362 | (0.49) | 1,169 | (0.70) | 2,901 | (0.64) |
| PB | 59 | (0.24) | 474 | (0.61) | 265 | (0.41) | 229 | (0.50) | 250 | (0.33) | 967 | (0.58) | 2,244 | (0.49) |
| PE | 292 | (1.21) | 733 | (0.94) | 225 | (0.34) | 178 | (0.39) | 240 | (0.32) | 408 | (0.24) | 2,076 | (0.46) |
| PI | 24 | (0.10) | 327 | (0.42) | 270 | (0.41) | 218 | (0.48) | 194 | (0.26) | 466 | (0.28) | 1,499 | (0.33) |
| PR | 105 | (0.43) | 879 | (1.13) | 1,446 | (2.22) | 1,151 | (2.53) | 1,614 | (2.16) | 4,050 | (2.43) | 9,245 | (2.03) |
| RJ | 1,930 | (7.98) | 2,748 | (3.52) | 1,798 | (2.76) | 1,989 | (4.38) | 2,232 | (2.99) | 3,966 | (2.38) | 14,663 | (3.23) |
| RN | 63 | (0.26) | 407 | (0.52) | 169 | (0.26) | 120 | (0.26) | 259 | (0.35) | 905 | (0.54) | 1,923 | (0.42) |
| RO | 26 | (0.11) | 312 | (0.40) | 323 | (0.49) | 206 | (0.45) | 369 | (0.49) | 748 | (0.45) | 1,984 | (0.44) |
| RR | 8 | (0.03) | 66 | (0.08) | 22 | (0.03) | 35 | (0.08) | 60 | (0.08) | 141 | (0.08) | 332 | (0.07) |
| RS | 104 | (0.43) | 691 | (0.89) | 1,400 | (2.15) | 1,231 | (2.71) | 1,504 | (2.02) | 5,098 | (3.05) | 10,028 | (2.21) |
| SC | 77 | (0.32) | 448 | (0.57) | 787 | (1.21) | 639 | (1.41) | 890 | (1.19) | 2,433 | (1.46) | 5,274 | (1.16) |
| SE | 25 | (0.10) | 256 | (0.33) | 153 | (0.23) | 80 | (0.18) | 189 | (0.25) | 472 | (0.28) | 1,175 | (0.26) |
| SP | 3,403 | (14.07) | 7,957 | (10.20) | 6,176 | (9.46) | 4,102 | (9.03) | 7,673 | (10.28) | 17,766 | (10.64) | 47,077 | (10.36) |
| TO | 5 | (0.02) | 56 | (0.07) | 101 | (0.15) | 53 | (0.12) | 77 | (0.10) | 237 | (0.14) | 529 | (0.12) |
| **no ICU** | **16,088** | **(66.53)** | **54,342** | **(69.65)** | **45,074** | **(69.07)** | **30,676** | **(67.52)** | **50,716** | **(67.96)** | **110,388** | **(66.12)** | **307,284** | **(67.61)** |
| AC | 2 | (0.01) | 138 | (0.18) | 72 | (0.11) | 24 | (0.05) | 51 | (0.07) | 88 | (0.05) | 375 | (0.08) |
| AL | 106 | (0.44) | 709 | (0.91) | 171 | (0.26) | 146 | (0.32) | 463 | (0.62) | 941 | (0.56) | 2,536 | (0.56) |
| AM | 1,059 | (4.38) | 1,666 | (2.14) | 835 | (1.28) | 1,026 | (2.26) | 3,631 | (4.87) | 1,667 | (1.00) | 9,884 | (2.17) |
| AP | 37 | (0.15) | 201 | (0.26) | 96 | (0.15) | 269 | (0.59) | 249 | (0.33) | 336 | (0.20) | 1,188 | (0.26) |
| BA | 190 | (0.79) | 1,844 | (2.36) | 1,292 | (1.98) | 819 | (1.80) | 1,495 | (2.00) | 3,364 | (2.01) | 9,004 | (1.98) |
| CE | 965 | (3.99) | 2,900 | (3.72) | 837 | (1.28) | 570 | (1.25) | 877 | (1.18) | 3,799 | (2.28) | 9,948 | (2.19) |
| DF | 106 | (0.44) | 1,711 | (2.19) | 2,702 | (4.14) | 900 | (1.98) | 856 | (1.15) | 2,875 | (1.72) | 9,150 | (2.01) |
| ES | 44 | (0.18) | 367 | (0.47) | 175 | (0.27) | 105 | (0.23) | 160 | (0.21) | 392 | (0.23) | 1,243 | (0.27) |
| GO | 75 | (0.31) | 1,145 | (1.47) | 2,532 | (3.88) | 1,123 | (2.47) | 1,401 | (1.88) | 4,414 | (2.64) | 10,690 | (2.35) |
| MA | 312 | (1.29) | 499 | (0.64) | 219 | (0.34) | 85 | (0.19) | 189 | (0.25) | 784 | (0.47) | 2,088 | (0.46) |
| MG | 229 | (0.95) | 3,095 | (3.97) | 4,167 | (6.39) | 2,642 | (5.82) | 5,252 | (7.04) | 11,738 | (7.03) | 27,123 | (5.97) |
| MS | 30 | (0.12) | 523 | (0.67) | 1,336 | (2.05) | 947 | (2.08) | 1,526 | (2.04) | 2,337 | (1.40) | 6,699 | (1.47) |
| MT | 63 | (0.26) | 1,153 | (1.48) | 1,086 | (1.66) | 414 | (0.91) | 962 | (1.29) | 1,679 | (1.01) | 5,357 | (1.18) |
| PA | 1,135 | (4.69) | 2,976 | (3.81) | 1,142 | (1.75) | 632 | (1.39) | 1,318 | (1.77) | 2,911 | (1.74) | 10,114 | (2.23) |
| PB | 134 | (0.55) | 1,130 | (1.45) | 665 | (1.02) | 539 | (1.19) | 701 | (0.94) | 1,692 | (1.01) | 4,861 | (1.07) |
| PE | 727 | (3.01) | 1,702 | (2.18) | 666 | (1.02) | 346 | (0.76) | 308 | (0.41) | 660 | (0.40) | 4,409 | (0.97) |
| PI | 104 | (0.43) | 1,263 | (1.62) | 763 | (1.17) | 564 | (1.24) | 522 | (0.70) | 1,253 | (0.75) | 4,469 | (0.98) |
| PR | 171 | (0.71) | 1,996 | (2.56) | 3,451 | (5.29) | 2,851 | (6.28) | 4,499 | (6.03) | 8,698 | (5.21) | 21,666 | (4.77) |
| RJ | 2,378 | (9.83) | 3,965 | (5.08) | 2,204 | (3.38) | 2,446 | (5.38) | 3,036 | (4.07) | 4,770 | (2.86) | 18,799 | (4.14) |
| RN | 87 | (0.36) | 974 | (1.25) | 285 | (0.44) | 190 | (0.42) | 489 | (0.66) | 1,335 | (0.80) | 3,360 | (0.74) |
| RO | 60 | (0.25) | 432 | (0.55) | 358 | (0.55) | 161 | (0.35) | 329 | (0.44) | 889 | (0.53) | 2,229 | (0.49) |
| RR | 14 | (0.06) | 274 | (0.35) | 101 | (0.15) | 109 | (0.24) | 221 | (0.30) | 153 | (0.09) | 872 | (0.19) |
| RS | 329 | (1.36) | 1,560 | (2.00) | 3,108 | (4.76) | 2,495 | (5.49) | 3,124 | (4.19) | 10,693 | (6.40) | 21,309 | (4.69) |
| SC | 146 | (0.60) | 924 | (1.18) | 2,050 | (3.14) | 1,880 | (4.14) | 2,423 | (3.25) | 6,014 | (3.60) | 13,437 | (2.96) |
| SE | 28 | (0.12) | 643 | (0.82) | 511 | (0.78) | 207 | (0.46) | 559 | (0.75) | 1,120 | (0.67) | 3,068 | (0.68) |
| SP | 7,534 | (31.16) | 20,214 | (25.91) | 13,760 | (21.08) | 9,022 | (19.86) | 15,874 | (21.27) | 35,260 | (21.12) | 101,664 | (22.37) |
| TO | 23 | (0.10) | 338 | (0.43) | 490 | (0.75) | 164 | (0.36) | 201 | (0.27) | 526 | (0.32) | 1,742 | (0.38) |

**Table S3.** Number of IMV requirements among COVID-19 patients aged 20-59 according to the Brazilian federative unit and outbreak periods. **'all'** accounts for hospitalized patients owning ventilation information. **'no IMV'** accounts for hospitalized patients requiring Non-Invasive Ventilation (NIV) or did not require ventilation.

| | T1 | (%) | T2 | (%) | T3 | (%) | T4 | (%) | T5 | (%) | T6 | (%) | Total | (%) |
|---|---|---|---|---|---|---|---|---|---|---|---|---|---|---|
| **all** | **22,387** | **(100.00)** | **74,263** | **(100.00)** | **62,691** | **(100.00)** | **43,712** | **(100.00)** | **72,095** | **(100.00)** | **164,204** | **(100.00)** | **439,352** | **(100.00)** |
| **IMV** | **4,146** | **(18.52)** | **11,982** | **(16.13)** | **8,978** | **(14.32)** | **5,527** | **(12.64)** | **10,441** | **(14.48)** | **33,506** | **(20.41)** | **74,580** | **(16.97)** |
| AC | 0 | (0.00) | 10 | (0.01) | 4 | (0.01) | 0 | (0.00) | 2 | (0.00) | 41 | (0.02) | 57 | (0.01) |
| AL | 72 | (0.32) | 385 | (0.52) | 57 | (0.09) | 34 | (0.08) | 82 | (0.11) | 148 | (0.09) | 778 | (0.18) |
| AM | 314 | (1.40) | 221 | (0.30) | 93 | (0.15) | 155 | (0.35) | 1,024 | (1.42) | 444 | (0.27) | 2,251 | (0.51) |
| AP | 17 | (0.08) | 97 | (0.13) | 16 | (0.03) | 40 | (0.09) | 74 | (0.10) | 203 | (0.12) | 447 | (0.10) |
| BA | 93 | (0.42) | 686 | (0.92) | 489 | (0.78) | 262 | (0.60) | 403 | (0.56) | 1,188 | (0.72) | 3,121 | (0.71) |
| CE | 392 | (1.75) | 740 | (1.00) | 236 | (0.38) | 143 | (0.33) | 270 | (0.37) | 1,382 | (0.84) | 3,163 | (0.72) |
| DF | 38 | (0.17) | 456 | (0.61) | 489 | (0.78) | 118 | (0.27) | 178 | (0.25) | 1,139 | (0.69) | 2,418 | (0.55) |
| ES | 55 | (0.25) | 276 | (0.37) | 91 | (0.15) | 101 | (0.23) | 116 | (0.16) | 324 | (0.20) | 963 | (0.22) |
| GO | 25 | (0.11) | 304 | (0.41) | 672 | (1.07) | 202 | (0.46) | 286 | (0.40) | 1,497 | (0.91) | 2,986 | (0.68) |
| MA | 64 | (0.29) | 112 | (0.15) | 40 | (0.06) | 21 | (0.05) | 52 | (0.07) | 281 | (0.17) | 570 | (0.13) |
| MG | 54 | (0.24) | 516 | (0.69) | 691 | (1.10) | 421 | (0.96) | 938 | (1.30) | 2,774 | (1.69) | 5,394 | (1.23) |
| MS | 5 | (0.02) | 56 | (0.08) | 205 | (0.33) | 114 | (0.26) | 267 | (0.37) | 716 | (0.44) | 1,363 | (0.31) |
| MT | 9 | (0.04) | 231 | (0.31) | 186 | (0.30) | 56 | (0.13) | 90 | (0.12) | 306 | (0.19) | 878 | (0.20) |
| PA | 225 | (1.01) | 430 | (0.58) | 131 | (0.21) | 102 | (0.23) | 239 | (0.33) | 857 | (0.52) | 1,984 | (0.45) |
| PB | 37 | (0.17) | 296 | (0.40) | 150 | (0.24) | 111 | (0.25) | 128 | (0.18) | 579 | (0.35) | 1,301 | (0.30) |
| PE | 170 | (0.76) | 416 | (0.56) | 88 | (0.14) | 64 | (0.15) | 67 | (0.09) | 150 | (0.09) | 955 | (0.22) |
| PI | 17 | (0.08) | 230 | (0.31) | 141 | (0.22) | 97 | (0.22) | 87 | (0.12) | 266 | (0.16) | 838 | (0.19) |
| PR | 53 | (0.24) | 384 | (0.52) | 656 | (1.05) | 503 | (1.15) | 762 | (1.06) | 2,724 | (1.66) | 5,082 | (1.16) |
| RJ | 789 | (3.52) | 1,076 | (1.45) | 509 | (0.81) | 492 | (1.13) | 723 | (1.00) | 1,300 | (0.79) | 4,889 | (1.11) |
| RN | 40 | (0.18) | 280 | (0.38) | 73 | (0.12) | 51 | (0.12) | 111 | (0.15) | 450 | (0.27) | 1,005 | (0.23) |
| RO | 17 | (0.08) | 237 | (0.32) | 173 | (0.28) | 89 | (0.20) | 261 | (0.36) | 566 | (0.34) | 1,343 | (0.31) |
| RR | 8 | (0.04) | 94 | (0.13) | 25 | (0.04) | 38 | (0.09) | 86 | (0.12) | 170 | (0.10) | 421 | (0.10) |
| RS | 55 | (0.25) | 414 | (0.56) | 797 | (1.27) | 599 | (1.37) | 812 | (1.13) | 3,792 | (2.31) | 6,469 | (1.47) |
| SC | 44 | (0.20) | 250 | (0.34) | 427 | (0.68) | 354 | (0.81) | 502 | (0.70) | 2,006 | (1.22) | 3,583 | (0.82) |
| SE | 14 | (0.06) | 278 | (0.37) | 148 | (0.24) | 86 | (0.20) | 197 | (0.27) | 477 | (0.29) | 1,200 | (0.27) |
| SP | 1,535 | (6.86) | 3,453 | (4.65) | 2,294 | (3.66) | 1,250 | (2.86) | 2,646 | (3.67) | 9,584 | (5.84) | 20,762 | (4.73) |
| TO | 4 | (0.02) | 54 | (0.07) | 97 | (0.15) | 24 | (0.05) | 38 | (0.05) | 142 | (0.09) | 359 | (0.08) |
| **no IMV** | **18,241** | **(81.48)** | **62,281** | **(83.87)** | **53,713** | **(85.68)** | **38,185** | **(87.36)** | **61,654** | **(85.52)** | **130,698** | **(79.59)** | **364,772** | **(83.03)** |
| AC | 1 | (0.00) | 128 | (0.17) | 61 | (0.10) | 25 | (0.06) | 21 | (0.03) | 71 | (0.04) | 307 | (0.07) |
| AL | 109 | (0.49) | 728 | (0.98) | 196 | (0.31) | 157 | (0.36) | 545 | (0.76) | 1,175 | (0.72) | 2,910 | (0.66) |
| AM | 1,012 | (4.52) | 1,626 | (2.19) | 846 | (1.35) | 1,032 | (2.36) | 3,550 | (4.92) | 1,759 | (1.07) | 9,825 | (2.24) |
| AP | 34 | (0.15) | 221 | (0.30) | 114 | (0.18) | 293 | (0.67) | 267 | (0.37) | 376 | (0.23) | 1,305 | (0.30) |
| BA | 247 | (1.10) | 2,377 | (3.20) | 1,758 | (2.80) | 1,318 | (3.02) | 2,193 | (3.04) | 4,675 | (2.85) | 12,568 | (2.86) |
| CE | 1,046 | (4.67) | 3,284 | (4.42) | 960 | (1.53) | 692 | (1.58) | 1,081 | (1.50) | 4,321 | (2.63) | 11,384 | (2.59) |
| DF | 132 | (0.59) | 2,364 | (3.18) | 3,190 | (5.09) | 1,173 | (2.68) | 1,203 | (1.67) | 3,580 | (2.18) | 11,642 | (2.65) |
| ES | 97 | (0.43) | 500 | (0.67) | 266 | (0.42) | 248 | (0.57) | 327 | (0.45) | 700 | (0.43) | 2,138 | (0.49) |
| GO | 92 | (0.41) | 1,355 | (1.82) | 2,961 | (4.72) | 1,434 | (3.28) | 1,920 | (2.66) | 5,374 | (3.27) | 13,136 | (2.99) |
| MA | 239 | (1.07) | 429 | (0.58) | 241 | (0.38) | 112 | (0.26) | 255 | (0.35) | 1,034 | (0.63) | 2,310 | (0.53) |
| MG | 294 | (1.31) | 3,460 | (4.66) | 4,755 | (7.58) | 3,201 | (7.32) | 6,101 | (8.46) | 13,738 | (8.37) | 31,549 | (7.18) |
| MS | 32 | (0.14) | 555 | (0.75) | 1,364 | (2.18) | 977 | (2.24) | 1,555 | (2.16) | 2,372 | (1.44) | 6,855 | (1.56) |
| MT | 82 | (0.37) | 1,220 | (1.64) | 1,152 | (1.84) | 528 | (1.21) | 1,234 | (1.71) | 1,770 | (1.08) | 5,986 | (1.36) |
| PA | 1,033 | (4.61) | 2,953 | (3.98) | 1,195 | (1.91) | 685 | (1.57) | 1,410 | (1.96) | 3,155 | (1.92) | 10,431 | (2.37) |
| PB | 137 | (0.61) | 1,244 | (1.68) | 762 | (1.22) | 646 | (1.48) | 811 | (1.12) | 1,978 | (1.20) | 5,578 | (1.27) |
| PE | 776 | (3.47) | 1,778 | (2.39) | 660 | (1.05) | 379 | (0.87) | 380 | (0.53) | 779 | (0.47) | 4,752 | (1.08) |
| PI | 101 | (0.45) | 1,338 | (1.80) | 889 | (1.42) | 705 | (1.61) | 631 | (0.88) | 1,564 | (0.95) | 5,228 | (1.19) |
| PR | 216 | (0.96) | 2,428 | (3.27) | 4,170 | (6.65) | 3,484 | (7.97) | 5,252 | (7.28) | 9,707 | (5.91) | 25,257 | (5.75) |
| RJ | 3,140 | (14.03) | 5,221 | (7.03) | 3,260 | (5.20) | 3,689 | (8.44) | 4,406 | (6.11) | 7,307 | (4.45) | 27,023 | (6.15) |
| RN | 108 | (0.48) | 1,049 | (1.41) | 360 | (0.57) | 261 | (0.60) | 653 | (0.91) | 1,777 | (1.08) | 4,208 | (0.96) |
| RO | 65 | (0.29) | 501 | (0.67) | 524 | (0.84) | 299 | (0.68) | 549 | (0.76) | 1,189 | (0.72) | 3,127 | (0.71) |
| RR | 14 | (0.06) | 218 | (0.29) | 90 | (0.14) | 108 | (0.25) | 203 | (0.28) | 117 | (0.07) | 750 | (0.17) |
| RS | 358 | (1.60) | 1,782 | (2.40) | 3,652 | (5.83) | 3,060 | (7.00) | 3,806 | (5.28) | 12,270 | (7.47) | 24,928 | (5.67) |
| SC | 169 | (0.75) | 1,039 | (1.40) | 2,304 | (3.68) | 2,101 | (4.81) | 2,713 | (3.76) | 6,258 | (3.81) | 14,584 | (3.32) |
| SE | 37 | (0.17) | 607 | (0.82) | 514 | (0.82) | 178 | (0.41) | 499 | (0.69) | 1,092 | (0.67) | 2,927 | (0.67) |
| SP | 8,648 | (38.63) | 23,552 | (31.71) | 17,000 | (27.12) | 11,222 | (25.67) | 19,853 | (27.54) | 42,016 | (25.59) | 122,291 | (27.83) |
| TO | 22 | (0.10) | 324 | (0.44) | 469 | (0.75) | 178 | (0.41) | 236 | (0.33) | 544 | (0.33) | 1,773 | (0.40) |

**Table S4.** Number of deaths among COVID-19 patients aged 20-59 according to the Brazilian federative unit and outbreak periods. **'all'** accounts for hospitalized patients with closed outcome (cure or death).

| | T1 | (%) | T2 | (%) | T3 | (%) | T4 | (%) | T5 | (%) | T6 | (%) | Total | (%) |
|---|---|---|---|---|---|---|---|---|---|---|---|---|---|---|
| **all** | **26,896** | **(100.00)** | **82,244** | **(100.00)** | **65,525** | **(100.00)** | **44,155** | **(100.00)** | **72,958** | **(100.00)** | **150,104** | **(100.00)** | **441,882** | **(100.00)** |
| **death** | **5,752** | **(21.39)** | **16,752** | **(20.37)** | **11,080** | **(16.91)** | **6,519** | **(14.76)** | **13,892** | **(19.04)** | **42,999** | **(28.65)** | **96,994** | **(21.95)** |
| AC | 7 | (0.03) | 86 | (0.10) | 22 | (0.03) | 15 | (0.03) | 43 | (0.06) | 151 | (0.10) | 324 | (0.07) |
| AL | 63 | (0.23) | 479 | (0.58) | 76 | (0.12) | 44 | (0.10) | 147 | (0.20) | 279 | (0.19) | 1,088 | (0.25) |
| AM | 483 | (1.80) | 354 | (0.43) | 147 | (0.22) | 209 | (0.47) | 1,877 | (2.57) | 674 | (0.45) | 3,744 | (0.85) |
| AP | 31 | (0.12) | 118 | (0.14) | 20 | (0.03) | 38 | (0.09) | 57 | (0.08) | 175 | (0.12) | 439 | (0.10) |
| BA | 99 | (0.37) | 825 | (1.00) | 619 | (0.94) | 311 | (0.70) | 487 | (0.67) | 1,291 | (0.86) | 3,632 | (0.82) |
| CE | 460 | (1.71) | 1,079 | (1.31) | 273 | (0.42) | 147 | (0.33) | 336 | (0.46) | 1,853 | (1.23) | 4,148 | (0.94) |
| DF | 17 | (0.06) | 412 | (0.50) | 484 | (0.74) | 119 | (0.27) | 138 | (0.19) | 1,069 | (0.71) | 2,239 | (0.51) |
| ES | 65 | (0.24) | 419 | (0.51) | 136 | (0.21) | 109 | (0.25) | 144 | (0.20) | 368 | (0.25) | 1,241 | (0.28) |
| GO | 22 | (0.08) | 421 | (0.51) | 858 | (1.31) | 296 | (0.67) | 376 | (0.52) | 2,378 | (1.58) | 4,351 | (0.98) |
| MA | 173 | (0.64) | 297 | (0.36) | 100 | (0.15) | 46 | (0.10) | 91 | (0.12) | 516 | (0.34) | 1,223 | (0.28) |
| MG | 36 | (0.13) | 670 | (0.81) | 916 | (1.40) | 569 | (1.29) | 1,301 | (1.78) | 4,463 | (2.97) | 7,955 | (1.80) |
| MS | 2 | (0.01) | 67 | (0.08) | 242 | (0.37) | 125 | (0.28) | 313 | (0.43) | 903 | (0.60) | 1,652 | (0.37) |
| MT | 10 | (0.04) | 314 | (0.38) | 301 | (0.46) | 78 | (0.18) | 164 | (0.22) | 410 | (0.27) | 1,277 | (0.29) |
| PA | 448 | (1.67) | 925 | (1.12) | 209 | (0.32) | 128 | (0.29) | 396 | (0.54) | 1,335 | (0.89) | 3,441 | (0.78) |
| PB | 62 | (0.23) | 438 | (0.53) | 190 | (0.29) | 133 | (0.30) | 161 | (0.22) | 780 | (0.52) | 1,764 | (0.40) |
| PE | 479 | (1.78) | 1,094 | (1.33) | 328 | (0.50) | 204 | (0.46) | 286 | (0.39) | 398 | (0.27) | 2,789 | (0.63) |
| PI | 16 | (0.06) | 260 | (0.32) | 164 | (0.25) | 104 | (0.24) | 83 | (0.11) | 299 | (0.20) | 926 | (0.21) |
| PR | 41 | (0.15) | 407 | (0.49) | 682 | (1.04) | 463 | (1.05) | 861 | (1.18) | 2,846 | (1.90) | 5,300 | (1.20) |
| RJ | 1,427 | (5.31) | 2,126 | (2.58) | 965 | (1.47) | 984 | (2.23) | 1,522 | (2.09) | 2,795 | (1.86) | 9,819 | (2.22) |
| RN | 39 | (0.15) | 377 | (0.46) | 86 | (0.13) | 58 | (0.13) | 160 | (0.22) | 562 | (0.37) | 1,282 | (0.29) |
| RO | 19 | (0.07) | 261 | (0.32) | 156 | (0.24) | 69 | (0.16) | 241 | (0.33) | 658 | (0.44) | 1,404 | (0.32) |
| RR | 7 | (0.03) | 123 | (0.15) | 26 | (0.04) | 29 | (0.07) | 82 | (0.11) | 198 | (0.13) | 465 | (0.11) |
| RS | 18 | (0.07) | 320 | (0.39) | 631 | (0.96) | 464 | (1.05) | 729 | (1.00) | 3,982 | (2.65) | 6,144 | (1.39) |
| SC | 27 | (0.10) | 179 | (0.22) | 384 | (0.59) | 269 | (0.61) | 479 | (0.66) | 1,973 | (1.31) | 3,311 | (0.75) |
| SE | 19 | (0.07) | 361 | (0.44) | 180 | (0.27) | 58 | (0.13) | 153 | (0.21) | 392 | (0.26) | 1,163 | (0.26) |
| SP | 1,676 | (6.23) | 4,266 | (5.19) | 2,752 | (4.20) | 1,411 | (3.20) | 3,219 | (4.41) | 12,007 | (8.00) | 25,331 | (5.73) |
| TO | 6 | (0.02) | 74 | (0.09) | 133 | (0.20) | 39 | (0.09) | 46 | (0.06) | 244 | (0.16) | 542 | (0.12) |
| **cure** | **21,144** | **(78.61)** | **65,492** | **(79.63)** | **54,445** | **(83.09)** | **37,636** | **(85.24)** | **59,066** | **(80.96)** | **107,105** | **(71.35)** | **344,888** | **(78.05)** |
| AC | 38 | (0.14) | 221 | (0.27) | 120 | (0.18) | 155 | (0.35) | 239 | (0.33) | 248 | (0.17) | 1,021 | (0.23) |
| AL | 142 | (0.53) | 938 | (1.14) | 235 | (0.36) | 195 | (0.44) | 609 | (0.83) | 1,068 | (0.71) | 3,187 | (0.72) |
| AM | 1,078 | (4.01) | 1,842 | (2.24) | 886 | (1.35) | 1,097 | (2.48) | 3,338 | (4.58) | 1,763 | (1.17) | 10,004 | (2.26) |
| AP | 25 | (0.09) | 212 | (0.26) | 112 | (0.17) | 295 | (0.67) | 281 | (0.39) | 399 | (0.27) | 1,324 | (0.30) |
| BA | 235 | (0.87) | 2,291 | (2.79) | 1,714 | (2.62) | 1,190 | (2.70) | 2,016 | (2.76) | 3,512 | (2.34) | 10,958 | (2.48) |
| CE | 1,103 | (4.10) | 3,270 | (3.98) | 986 | (1.50) | 694 | (1.57) | 1,105 | (1.51) | 3,532 | (2.35) | 10,690 | (2.42) |
| DF | 160 | (0.59) | 2,413 | (2.93) | 3,107 | (4.74) | 1,070 | (2.42) | 1,116 | (1.53) | 2,991 | (1.99) | 10,857 | (2.46) |
| ES | 100 | (0.37) | 521 | (0.63) | 263 | (0.40) | 252 | (0.57) | 265 | (0.36) | 447 | (0.30) | 1,848 | (0.42) |
| GO | 106 | (0.39) | 1,437 | (1.75) | 3,220 | (4.91) | 1,429 | (3.24) | 1,894 | (2.60) | 4,467 | (2.98) | 12,553 | (2.84) |
| MA | 462 | (1.72) | 641 | (0.78) | 207 | (0.32) | 100 | (0.23) | 262 | (0.36) | 892 | (0.59) | 2,564 | (0.58) |
| MG | 313 | (1.16) | 3,675 | (4.47) | 5,032 | (7.68) | 3,291 | (7.45) | 6,241 | (8.55) | 11,690 | (7.79) | 30,242 | (6.84) |
| MS | 47 | (0.17) | 595 | (0.72) | 1,489 | (2.27) | 1,096 | (2.48) | 1,537 | (2.11) | 1,541 | (1.03) | 6,305 | (1.43) |
| MT | 73 | (0.27) | 1,100 | (1.34) | 1,084 | (1.65) | 417 | (0.94) | 1,003 | (1.37) | 1,298 | (0.86) | 4,975 | (1.13) |
| PA | 1,090 | (4.05) | 2,743 | (3.34) | 1,069 | (1.63) | 645 | (1.46) | 1,253 | (1.72) | 2,368 | (1.58) | 9,168 | (2.07) |
| PB | 118 | (0.44) | 1,049 | (1.28) | 640 | (0.98) | 510 | (1.16) | 691 | (0.95) | 1,555 | (1.04) | 4,563 | (1.03) |
| PE | 1,935 | (7.19) | 3,177 | (3.86) | 1,073 | (1.64) | 564 | (1.28) | 556 | (0.76) | 805 | (0.54) | 8,110 | (1.84) |
| PI | 112 | (0.42) | 1,168 | (1.42) | 615 | (0.94) | 424 | (0.96) | 319 | (0.44) | 733 | (0.49) | 3,371 | (0.76) |
| PR | 235 | (0.87) | 2,339 | (2.84) | 4,112 | (6.28) | 3,419 | (7.74) | 4,954 | (6.79) | 8,092 | (5.39) | 23,151 | (5.24) |
| RJ | 3,083 | (11.46) | 4,950 | (6.02) | 2,645 | (4.04) | 2,851 | (6.46) | 3,917 | (5.37) | 5,179 | (3.45) | 22,625 | (5.12) |
| RN | 107 | (0.40) | 956 | (1.16) | 327 | (0.50) | 216 | (0.49) | 502 | (0.69) | 995 | (0.66) | 3,103 | (0.70) |
| RO | 64 | (0.24) | 576 | (0.70) | 668 | (1.02) | 306 | (0.69) | 481 | (0.66) | 642 | (0.43) | 2,737 | (0.62) |
| RR | 14 | (0.05) | 222 | (0.27) | 97 | (0.15) | 116 | (0.26) | 240 | (0.33) | 113 | (0.08) | 802 | (0.18) |
| RS | 419 | (1.56) | 1,964 | (2.39) | 3,898 | (5.95) | 3,212 | (7.27) | 3,603 | (4.94) | 9,659 | (6.43) | 22,755 | (5.15) |
| SC | 213 | (0.79) | 1,286 | (1.56) | 2,565 | (3.91) | 2,316 | (5.25) | 2,879 | (3.95) | 6,304 | (4.20) | 15,563 | (3.52) |
| SE | 34 | (0.13) | 336 | (0.41) | 174 | (0.27) | 119 | (0.27) | 200 | (0.27) | 474 | (0.32) | 1,337 | (0.30) |
| SP | 9,827 | (36.54) | 25,333 | (30.80) | 17,693 | (27.00) | 11,522 | (26.09) | 19,404 | (26.60) | 36,067 | (24.03) | 119,846 | (27.12) |
| TO | 11 | (0.04) | 237 | (0.29) | 414 | (0.63) | 135 | (0.31) | 161 | (0.22) | 271 | (0.18) | 1,229 | (0.28) |

**Table S5.** In-hospital fatality risk ratio of COVID-19 patients aged 20-59 years in the last wave vs. the first wave

| | Risk Ratio[a] | 95% CI | p-values |
|---|---|---|---|
| **Brasil (T6/T2)** | **1.41** | **[1.38,1.43]** | <0.001 |
| Amazonas (T5/T1) | 1.16 | [1.07,1.26] | <0.001 |
| Goiás (T6/T3) | 1.65 | [1.54,1.77] | <0.001 |
| Minas Gerais (T6/T3) | 1.79 | [1.68,1.91] | <0.001 |
| Paraná (T6/T3) | 1.83 | [1.69,1.97] | <0.001 |
| Rio de Janeiro (T6/T2) | 1.17 | [1.11,1.22] | <0.001 |
| Rio Grande do Sul (T6/T3) | 2.10 | [1.94,2.26] | <0.001 |
| Santa Catarina (T6/T3) | 1.83 | [1.65,2.03] | <0.001 |
| São Paulo (T6/T2) | 1.73 | [1.68,1.79] | <0.001 |

[a]Risk Ratio = (hCFR of the last wave)/(hCFR of the first wave). Last wave (T5 or T6) and first wave (T1, T2 or T3), according to the federative unit.
